# C5L2 Gene Polymorphisms and Their Interaction with Metabolic and Inflammatory Pathways in T2DM-Associated Coronary Heart Disease: Insights from an Integrative Genetic and Clinical Analysis

**DOI:** 10.1101/2025.05.04.25326947

**Authors:** Mengjia Liu, Fen Liu, Abulayti Diljhumal, Juan Yang, Yating Wang, Meng Ye, Taotao Jia, Ying Gao

## Abstract

This study investigates the role of C5L2 gene polymorphisms and their interaction with metabolic and inflammatory factors in the development of coronary heart disease (CHD) among individuals with type 2 diabetes mellitus (T2DM) in a Chinese population from Xinjiang. The C5L2 gene encodes a receptor involved in lipid metabolism, immune regulation, and inflammatory signaling. A total of 951 participants were recruited in a hospital-based case-control study, including 206 patients with comorbid T2DM and CHD and 745 age- and sex-matched healthy controls. Clinical and biochemical parameters were measured using standardized laboratory procedures. Two single-nucleotide polymorphisms (rs2972607 and rs8112962) in the C5L2 gene were genotyped using the improved multiplex ligation detection reaction method. Statistical analyses were conducted to evaluate associations between genotypes, clinical traits, and disease risk, and multifactor dimensionality reduction analysis was used to explore gene–environment interactions. Individuals in the case group showed significantly higher levels of glucose, triglyceride-glucose index, white blood cells, lactate dehydrogenase, and creatine kinase compared to controls. The rs2972607 variant was independently associated with increased risk of T2DM-associated CHD and was significantly correlated with HDL cholesterol, platelet indices, and liver function markers. Interaction analyses revealed complex gene–environment networks, with glucose and the TyG index emerging as central predictors. The rs2972607 polymorphism appeared to modulate the strength of these interactions, suggesting a mechanistic role in disease susceptibility. These findings highlight the value of integrative genetic and clinical analyses for improving risk stratification and support the potential utility of C5L2 genotyping in personalized prevention strategies for cardiometabolic disease.

## 1. Introduction

Coronary heart disease (CHD) is an ischemic condition arising from the narrowing or occlusion of the coronary arteries and remains the leading cause of cardiovascular disease-related mortality on a global scale. The World Heart Report 2023 indicates that cardiovascular diseases claimed approximately 20.5 million lives in 2021, accounting for nearly one-third of all deaths [1]. At the same time, the China Cardiovascular Health and Disease Report 2023 highlights that cardiovascular diseases continue to be the primary cause of death among both urban and rural populations in China, with an estimated 11.39 million CHD cases, thereby underscoring the profound challenges in disease prevention and control [2].

Diabetes mellitus (DM), particularly type 2 diabetes mellitus (T2DM), is closely linked to CHD through shared pathophysiological pathways. T2DM is predominantly characterized by insulin resistance, a condition that substantially elevates the risk of developing CHD [3]. Chronic hyperglycemia associated with T2DM causes a cascade of deleterious effects, most notably, vascular endothelial damage, oxidative stress, inflammation, and metabolic dysregulation, which collectively accelerate atherosclerosis and thereby heighten CHD risk [4].

Recent studies have brought attention to the role of Complement 5a receptor 2 (C5L2) in the regulation of inflammatory and metabolic processes., This receptor is posited to play a pivotal role in the pathophysiological continuum linking T2DM and CHD [5,6]. In addition, biomarkers like the platelet-lymphocyte ratio (PLR), the atherogenic index of plasma (AIP), and the triglyceride-glucose index (TyG) have emerged as valuable indicators of metabolic and inflammatory status. These indices correlate strongly with the progression of both T2DM and CHD, thereby offering promise in improving disease prediction and evaluation [7-9].

The development of CHD and T2DM is influenced by a variety of factors, including lifestyle choices, psychological stress, and genetic predisposition. However, research exploring the role of genetic variants and their contribution to the risk of these interrelated conditions is still limited. A seminal study conducted in 2017 identified a potential association between CHD and two single-nucleotide polymorphisms (SNPs), rs2972607 and rs8112962 within the C5L2 gene in a Chinese Han population [10]. Although these variants have been implicated in CHD, their exact role in the combined pathogenesis of T2DM and CHD has not been fully understood.

To address these gaps, the present study adopts a genetic-environment interaction (G×E) model to examine how polymorphisms at the C5L2 locus are associated with emerging cardiovascular markers (such as AIP and TyG) alongside traditional risk factors (including smoking, alcohol consumption, and dyslipidemia). By elucidating these interactions, our research aims to advance the understanding of the shared pathophysiological mechanisms underlying T2DM and CHD, ultimately providing a robust basis for the early detection and stratification of high-risk individuals.

## 2. Materials and Methods

### 2.1 Study population

This hospital-based case–control study matched participants by age and sex in a 1:3 ratio (cases: controls). Sample size was estimated using a literature-based prevalence (PL = 0.046), an expected odds ratio of 2.6, α = 0.05, and 90% power (β = 0.10), yielding a minimum requirement of 156 cases and 468 controls. The target sample was increased by 30% to reduce bias, resulting in 206 cases and 745 controls. From January 2021 to June 2024, 951 unrelated Chinese adults were consecutively recruited from the First Affiliated Hospital of Xinjiang Medical University. The case group included 206 patients (124 men, 82 women; mean age 58 ± 13 years) with both coronary heart disease (CHD) and type 2 diabetes mellitus (T2DM). The control group comprised 745 participants (494 men, 251 women; mean age 56 ± 15 years) without CHD or T2DM. All participants underwent coronary angiography, independently reviewed by two cardiologists. CHD was defined as ≥50% luminal stenosis in any major coronary artery or branch. T2DM diagnosis required clinical symptoms plus one: FPG ≥ 7.0 mmol/L, glucose ≥ 11.1 mmol/L, HbA1c ≥ 6.5%, random glucose ≥ 11.1 mmol/L, or documented history. Patients with aortic coarctation, severe heart failure, cardiogenic shock, malignant arrhythmias, renal/liver disease, endocrine or metabolic disorders, malignancies, or autoimmune diseases were excluded. All participants gave written informed consent. The study followed the Declaration of Helsinki and was approved by the Ethics Committee of the First Affiliated Hospital of Xinjiang Medical University (Approval No. 240424-14).

### 2.2. Blood sample collection and hematological analysis

All participants provided informed consent and completed a demographic questionnaire capturing age, sex, and smoking history. After an overnight fast (from 8 p.m.), 2 mL of venous blood was collected the next morning into EDTA tubes, centrifuged at 3000 rpm for 15 minutes, and the plasma was stored at –80L°C for further analysis. All assays were performed using standardized protocols at the Laboratory Center of Xinjiang Medical University’s First Affiliated Hospital. Coagulation parameters were measured using an automated coagulation analyzer (CS-5100, Sysmex, Japan), and routine hematological indices were assessed with an automated blood cell analyzer (CAL8000, Myriad, China). The clinical panel included RBC, Hb, WBC, lymphocytes (LY), monocytes (MONO), eosinophils (EOS), basophils (BASO), LDH, PCV, MCV, MCH, RDW, PLT, MPV, PT, and APTT. Biochemical markers included glucose, triglycerides (TG), total cholesterol (TC), HDL-C, LDL-C, apolipoprotein A (apo-A), apolipoprotein B (apo-B), lipoprotein(a), and blood urea nitrogen (BUN). Several derived indices were calculated: atherogenic index of plasma (AIP = log[TG/HDL-C]), triglyceride-glucose index (TyG = ln[(TG × FPG)/2]), systemic inflammatory response index (SIRI = NEUT × MONO/LY), systemic immune-inflammation index (SII = PLT × NEUT/LY), neutrophil-to-lymphocyte ratio (NLR = NEUT/LY), platelet-to-lymphocyte ratio (PLR = PLT/LY), and basophil-to-albumin ratio (BAR = BUN/albumin). This approach ensured a comprehensive evaluation of hematological and biochemical status using consistent, validated methods.

### 2.3 Genomic DNA Extraction and SNP Genotyping

Following hospital admission, 3 ml of peripheral venous blood was aseptically collected from each participant into EDTA-containing tubes. Leukocytes were promptly isolated, and genomic DNA was extracted using the standard phenol–chloroform method. Isolated leukocytes were stored at –80 °C for future analyses.

Two C5L2 SNPs (rs2972607 and rs8112962) were selected based on linkage disequilibrium (r²L≥L0.8) and minor allele frequency (MAFL≥L0.05), identified via Haploview 4.2 using data from the International HapMap Project (Phase I & II) [10]. Genotyping was performed using the improved Multiplex Ligation Detection Reaction (iMLDR) technique (Genesky Biotechnologies Inc., Shanghai, China). Primer sequences are provided in Supplementary Table 1. All procedures were blinded, and 10% of samples were randomly re-genotyped for quality control. Genotypes were classified as AA, GA, GG (rs2972607) and TT, CT, CC (rs8112962).

### 2.4. Statistical analysis

All statistical analyses were performed using SPSS 26.0. Normally distributed data are presented as mean ± SD and compared using independent samples t-tests; non-normally distributed data are shown as median (IQR) and analyzed via rank sum and Mann–Whitney tests. Categorical variables were summarized as n (%) and assessed using chi-square tests, which were also applied to evaluate genotypic and allelic distributions and Hardy–Weinberg equilibrium. Associations between C5L2 polymorphisms and disease risk were examined using logistic regression, with adjusted odds ratios (ORs) and 95% confidence intervals (CIs) reported. Forest plots of multivariable logistic regression were generated in GraphPad Prism 9.5.0. Gene–environment interactions were explored using multifactor dimensionality reduction (MDR 3.0.2). All tests were two-sided, with significance set at P < 0.05.

### 2.5. Methodological workflow

The study followed a structured workflow integrating clinical assessment, biochemical analysis, and genetic profiling to evaluate the interaction between *CETP* gene polymorphisms and environmental factors in patients with coronary heart disease (CHD) and type 2 diabetes mellitus (T2DM). Participants were recruited based on established inclusion and exclusion criteria. Clinical data (age, gender, smoking, alcohol consumption, and medical history) and biochemical markers (fasting blood sugar, lipid profile, and blood pressure) were systematically collected. Genomic DNA was isolated from peripheral leukocytes using a salting-out method and analyzed for *CETP* gene variants rs708272 and rs1800775 via PCR-based genotyping. The associations between genetic variants and clinical indicators were assessed, followed by evaluation of gene-environment interactions influencing CHD susceptibility in the context of T2DM. A detailed overview of the methodological workflow is presented in Figure 1.

**Figure 1.**
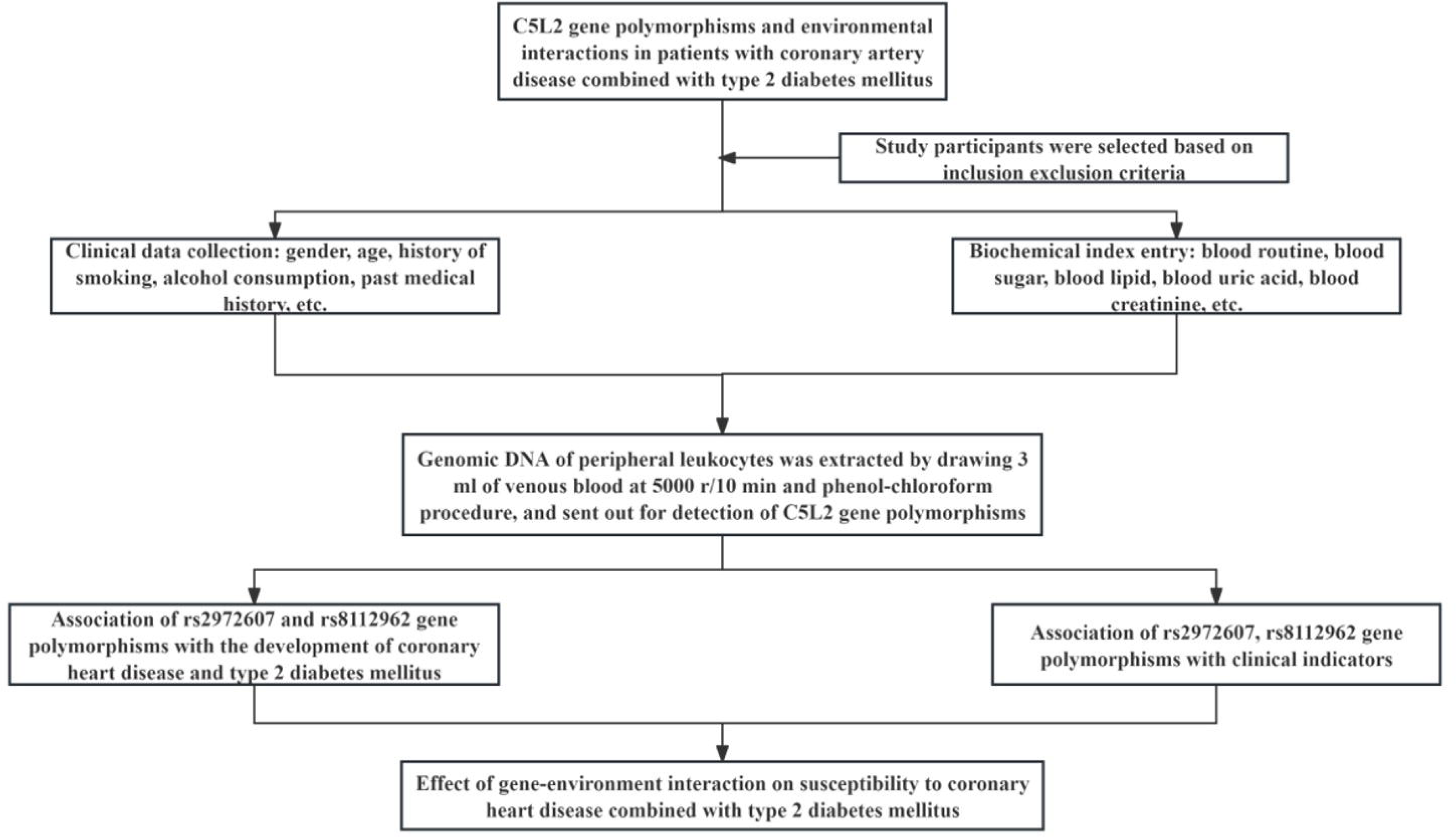
Study Design Workflow for Evaluating the Role of C5L2 Gene Polymorphisms and Gene–Environment Interactions in T2DM with CHD. Flowchart summarizing the study design. Participants with T2DM and CHD, along with healthy controls, underwent clinical assessment and genotyping for C5L2 polymorphisms (rs2972607, rs8112962). Associations with disease risk were evaluated through independent and gene–environment interaction analyses

## 3. Results

### 3.1. Baseline characteristics of study participants across clinical groups

A total of 951 participants were enrolled, including 206 cases and 745 controls. The case group (124 males, 82 females; mean age 58L±L13 years) was slightly older than the control group (494 males, 251 females; mean age 56L±L15 years). Comparative analysis revealed significant differences (PL<L0.05) in multiple biochemical and clinical parameters. Cases exhibited elevated levels of inflammatory and metabolic markers such as WBC, NEUT, MONO, BASO, NEp, BAp, LDH, CK, CK-MB, RBC, PT, BUN, glucose, GSP, TG, LP(a), UCB, globulin, AST, ALT, GGT, and 5′-NT. Derived indices, including AIP, SIRI, SII, TyG, NLR, BAR, and PLR, were also significantly higher, indicating heightened inflammation and metabolic dysfunction. Conversely, EOS, LYp, MOp, EOp, MCV, MCH, RDW, PDW, APTT, UA, HDL-C, Apo-A, CB, albumin, and A/G were significantly lower in the case group (PL<L0.05), suggesting altered hematological and hepatic profiles. No significant differences were observed in sex, ethnicity, alcohol/smoking history, Hb, or PCV (PL≥L0.05), indicating well-matched groups. A visual overview of significantly altered clinical markers is shown in Figure 2, with detailed characteristics provided in Supplementary Table 2.

**Figure 2.**
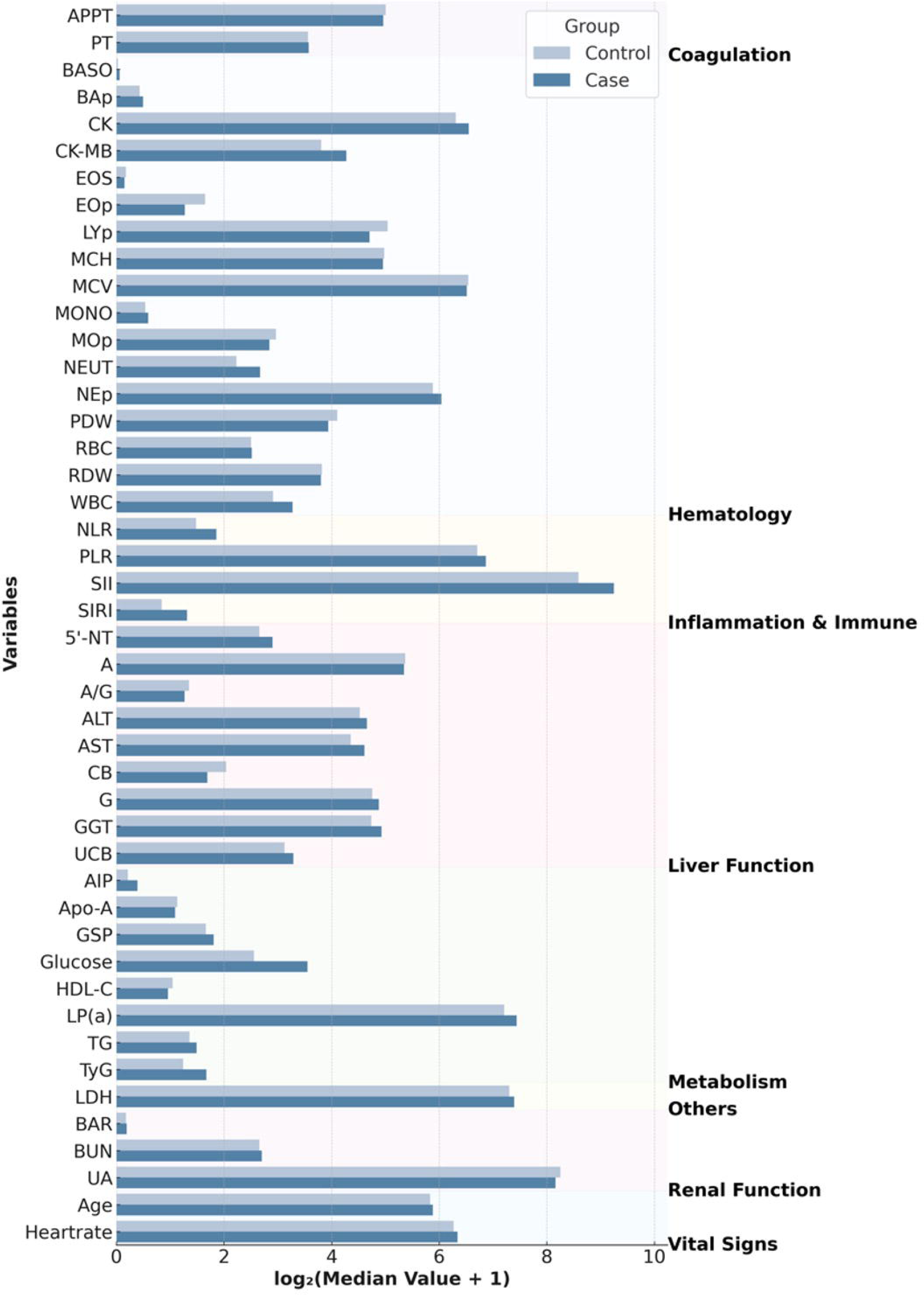
Comparison of Significantly Different Clinical Characteristics Between Case and Control Groups. Bar plot illustrates the median values of clinical parameters that showed statistically significant differences (P < 0.05) between the case and control groups. Bars are shaded in light blue for controls and dark blue for cases. Variables were categorized into seven physiological systems based on their primary biological role.

### 3.2. Association of rs2972607 and rs8112962 Polymorphisms with Susceptibility to T2DM with Coronary Heart Disease

Genotype distributions for both rs2972607 and rs8112962 polymorphisms conformed to Hardy-Weinberg equilibrium in cases and controls (P ≥ 0.05), indicating genetic equilibrium and representativeness of the study population (Supplementary Fig. 1, Supplementary Table 3). For rs2972607, the AA genotype was significantly underrepresented in the case group under the dominant model (AA vs. GG+GA, P = 0.017), and A allele frequency was reduced compared to controls (P = 0.050). Conversely, the GA genotype was significantly more frequent among cases in the overdominant model (GA vs. AA+GG, P = 0.009), as shown by the blue line in Fig. 3. These findings suggest a protective role for the AA genotype and potential heterozygote advantage contributing to G allele persistence through balancing selection. Similarly, for rs8112962, the TT genotype was less frequent in the case group under the dominant model (TT vs. CT+CC, P = 0.027), indicating a protective effect. The TC genotype was significantly enriched in cases under the overdominant model (TC vs. TT+CC, P = 0.016), visualized by the orange line in Fig. 3 (see also Supplementary Table 4). Taken together, these observations indicate that while the C allele may have deleterious effects in homozygosity while its persistence, particularly in heterozygous form, suggests possible selective advantage, incomplete penetrance, or genetic drift. Overall, these results validate the genetic representativeness of the study cohort and highlight the nuanced role of rs2972607 and rs8112962 variants in T2DM+CHD susceptibility, supporting the relevance of considering non-Mendelian inheritance patterns in complex disease genetics.

**Figure 3.**
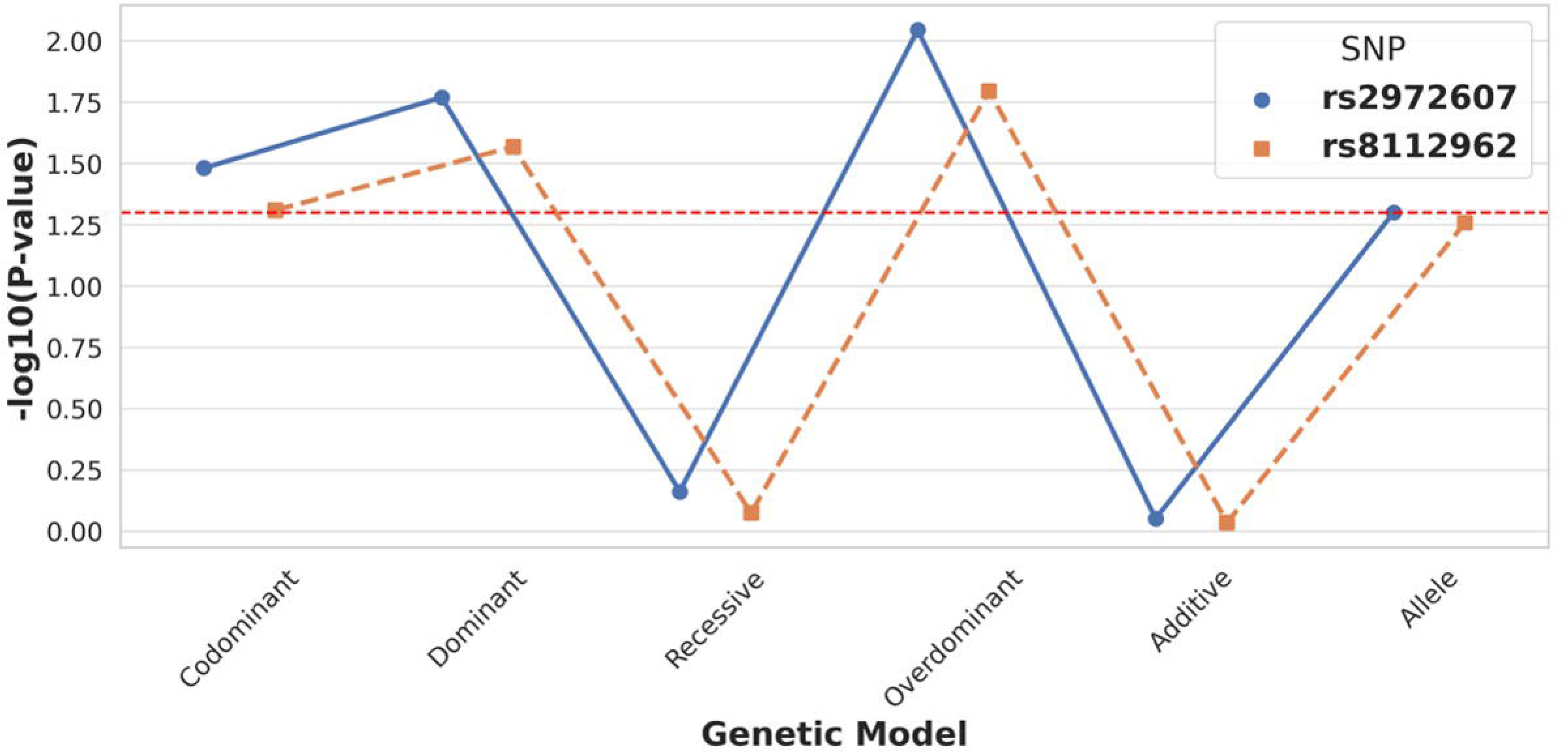
Comparative Analysis of Genotypic Models for rs2972607 and rs8112962 in Case. The figure displays logLL transformed P-values for six genetic models (codominant, dominant, recessive, overdominant, additive, and allelic) comparing C5L2 SNPs rs2972607 and rs8112962 between T2DM+CHD cases and controls. The red dashed line marks statistical significance (P = 0.05). Overdominant and dominant models, especially for rs2972607, show the strongest associations with disease risk.

### 3.3. Association of C5L2 rs2972607 and rs8112962 Genotypes with Clinical and Biochemical Indicators

At the rs2972607 locus of the C5L2 gene, three genotypic variants, AA (wild-type), GA, and GG (mutant types), were identified. Similarly, at the rs8112962 locus, three genotypes were observed: TT (wild-type), CT, and CC (mutant types). Analysis of clinical parameters across subgroups of the rs2972607 genotypes revealed statistically significant differences in several indicators including breathing rate (RR), high-density lipoprotein cholesterol (HDL-C), weight, platelet count (PLT), De Ritis ratio (AST/ALT), unconjugated bilirubin (UCB), 5’-nucleotidase (5’-NT), lymphocyte count (LY) and platelet distribution width levels (P < 0.05). These associations are visually summarized in Figure 4a, corresponding to Supplementary Table 5a. Similarly, for the rs8112962 genotypes, significant differences were observed in monocyte percentage (MOp), monocyte count (MONO), breathing rate (RR), and HDL-C across genotype subgroups (P < 0.05), as shown in Figure 4b and detailed in Supplementary Table 5b. Despite these genotype-specific associations with biochemical and hematological markers, no statistically significant associations were found in either rs2972607 or rs8112962 genotypes with age, gender, smoking, or alcohol consumption (Drinking) (P ≥ 0.05). Collectively, our results suggest that CSL2 gene polymorphisms exert their influence through direct biochemical or metabolic pathways, independent of common external lifestyle factors.

**Figure 4.**
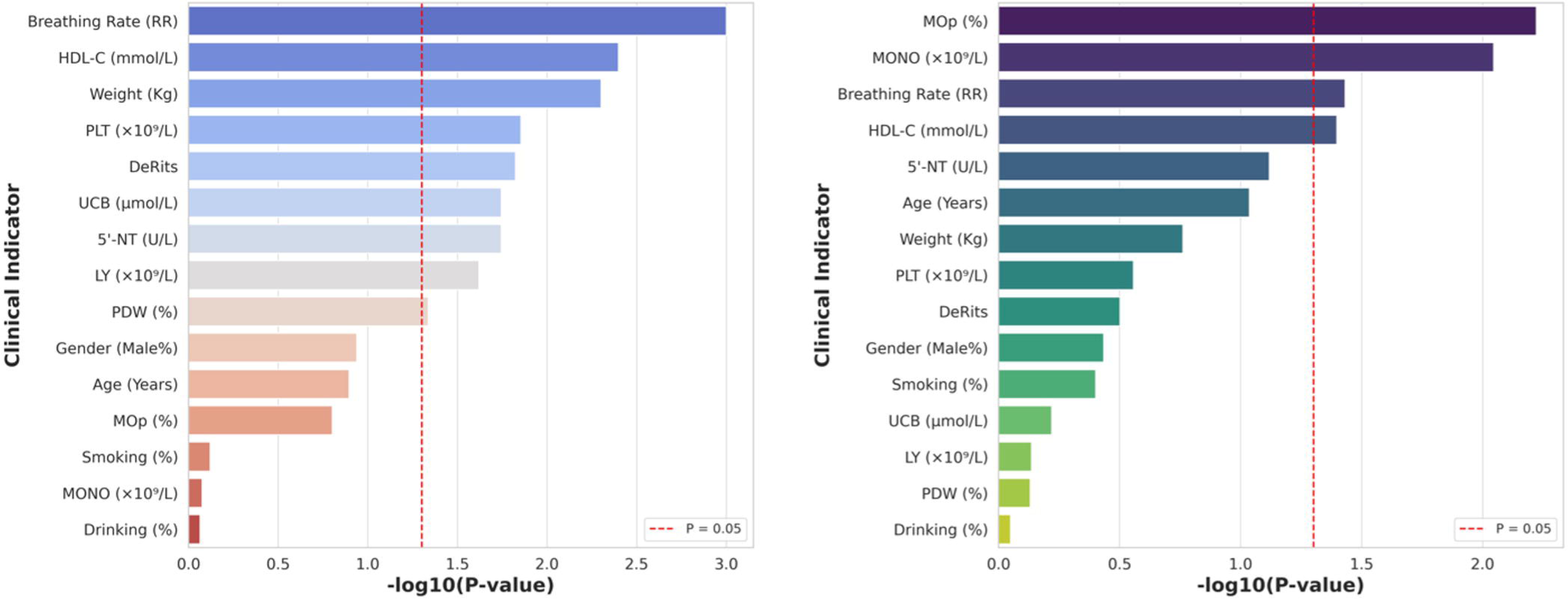
Associations Between C5L2 Genotypes (rs2972607 and rs8112962) and Clinical Indicators. Bar plots displaying –logLL transformed P-values for associations between clinical/biochemical indicators and C5L2 gene polymorphisms rs2972607 (A) and rs8112962 (B). The red dashed line (–logLL[P] = 1.3) marks the significance threshold (P < 0.05). For rs2972607, significant associations were observed with weight, respiratory rate, lymphocyte count, platelet indices, HDL-C, and liver markers. For rs8112962, HDL-C, monocyte count, monocyte percentage, and respiratory rate showed significant correlations.

### 3.4. Analysis of T2DM and CHD in the Chinese Population Using Logistic Regression

#### 3.4.1. Univariate analysis of clinical and genetic indicators

To assess the contribution of clinical and genetic factors to comorbid type 2 diabetes mellitus (T2DM) and coronary heart disease (CHD), we performed univariate logistic regression using variables categorized per the 2024 Adult Hyperuricemia and Gout Dietary Guidelines (Supplementary Table 6). As shown in Figure 5A and Supplementary Table 7, several physiological (respiratory rate, heart rate), hematological (WBC, MONO, LY, EOS, PLT), coagulation (APTT), metabolic (UA, fasting glucose, HDL-C, CB, UCB, GGT, 5’-NT), and systemic indices (AIP, SII, TyG, PLR) were significantly associated with T2DM+CHD (P < 0.05). Notably, the C5L2 polymorphism rs2972607 also showed a significant association (P = 0.034).

**Figure 5.**
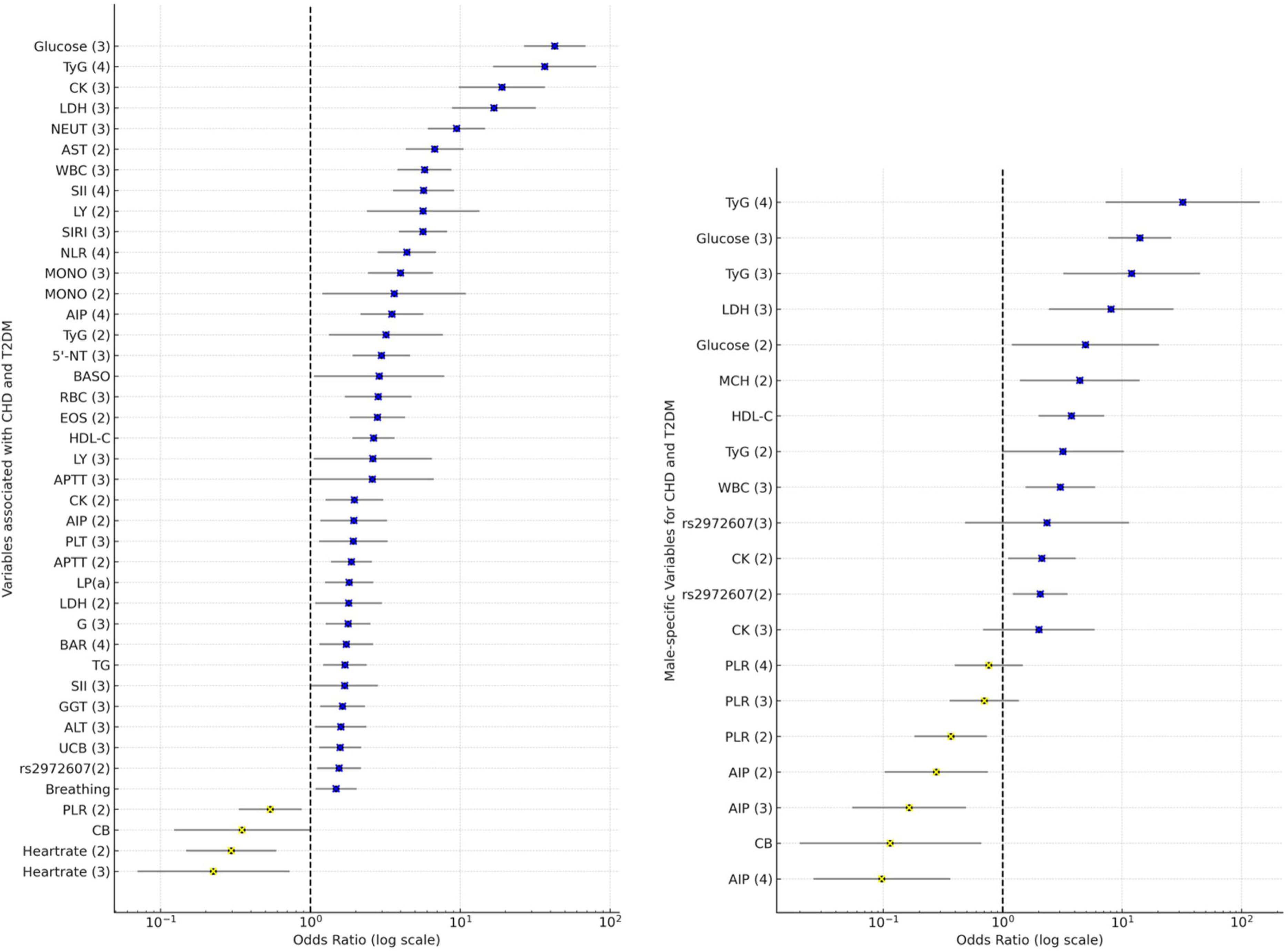
Predictors of T2DM with CHD Identified by (A) Univariate and (B) Multivariate Logistic Regression Analysis. Forest plots showing odds ratios (ORs) and 95% confidence intervals (CIs) for biochemical, genetic, and hematological variables associated with comorbid type 2 diabetes mellitus (T2DM) and coronary heart disease (CHD). The dashed line at OR = 1 denotes no effect. OR > 1 (blue) indicates increased risk; OR < 1 (yellow) indicates protection. Univariate analysis highlights glucose, TyG index, CK, LDH, and NEUT as risk factors, while heart rate, PLR, and CB are protective. Similarly, multivariate analysis confirms independent associations for glucose, TyG, LDH, WBC, and rs2972607, with lower AIP and PLR maintaining protective effects.

#### 3.4.2. Multivariate analysis identifies independent predictors

To identify independent risk factors for T2DM+CHD, variables significant in univariate analysis were included in a multivariate logistic regression model (Figure 5B, Supplementary Table 8). Fasting glucose emerged as the strongest predictor (OR = 14.08; 95% CI: 7.67–25.87; P < 0.001), followed by WBC, MCH, HDL-C, CB, LDH, and CK. Inflammatory and metabolic markers (PLR, AIP) also remained significant, underscoring the role of systemic inflammation and metabolic dysregulation. Notably, the C5L2 rs2972607 genotype remained independently associated with increased risk (OR = 2.07; 95% CI: 1.22–3.50; P = 0.007), supporting its potential as a genetic susceptibility marker. These findings highlight the combined contribution of clinical and genetic factors to T2DM+CHD pathogenesis and the value of integrated biomarker-genotype models for risk stratification.

### 3.5. The Relationship Between the *C5L2* Gene, Environmental Factors, and the Onset of T2DM and CHD

To investigate the interplay between genetic and environmental factors in the pathogenesis of type 2 diabetes mellitus (T2DM) and coronary heart disease (CHD), we used multifactor dimensionality reduction (MDR) to identify high-order interactions involving C5L2 polymorphisms (rs2972607, rs8112962) and clinical/biochemical variables, as shown in Figure 5A. The best model combined these SNPs with 14 factors like gender, smoking, drinking, age, WBC, MCH, glucose, LDH, AIP, TyG, PLR, HDL-C, CB, and CK, achieving perfect cross-validation (10/10), high significance (P < 0.001), and strong predictive metrics (training balanced accuracy = 0.9957; testing = 0.6057).

Glucose consistently appeared in all top models, underscoring its central role. Notably, adding rs2972607 improved interaction strength, while inclusion of rs8112962 revealed a synergistic effect, suggesting enhanced susceptibility through combined genetic and metabolic perturbations. Although glucose alone was the best single predictor (testing balanced accuracy = 0.8633), its power increased with AIP, TyG, PLR, and WBC, highlighting the disease’s multifactorial nature.

The integration of lipid indices (HDL-C, AIP) and inflammatory markers (WBC, PLR) with genetic data supports a model of immune-metabolic dysregulation driving disease risk. Overall, these findings reveal a robust, multifactorial interaction network, where C5L2 variants, especially rs2972607, interact with metabolic and inflammatory markers rather than acting independently. Figure 5B confirms the added value of genetic data in uncovering gene-environment interactions, while also illustrating the balance between model complexity and generalizability. This high-order model offers novel insights into T2DM with CHD and may guide future risk stratification and targeted interventions. The complete MDR model hierarchy and associated performance metrics are presented in Supplementary Table 10.

#### 3.5.1 Gene-Gene Interaction Analyses

Multifactor Dimensionality Reduction (MDR) analysis was used to assess potential epistatic interactions between the C5L2 polymorphisms rs2972607 and rs8112962 to comorbid type 2 diabetes mellitus (T2DM) and coronary heart disease (CHD). As shown in Figure 6B, genotype combinations were classified as high-risk (dark), low-risk (light), or neutral (white), with vertical bars representing case (left) and control (right) counts. Combinations such as TT-AA and CT-GA appeared more frequently among cases, indicating elevated risk. Figure 6C summarizes interaction effects using the information gain metric. A negative interaction information gain (–0.42%) suggests a slight redundancy, indicating that the SNPs contribute independently rather than synergistically to disease risk. Individually, rs2972607 and rs8112962 showed notable effects, with information gains of 0.50% and 0.44%, respectively, supporting their roles as independent genetic risk factors. In summary, while no strong interaction was detected, both SNPs independently contribute to CHD risk in individuals with T2DM.

**Figure 6.**
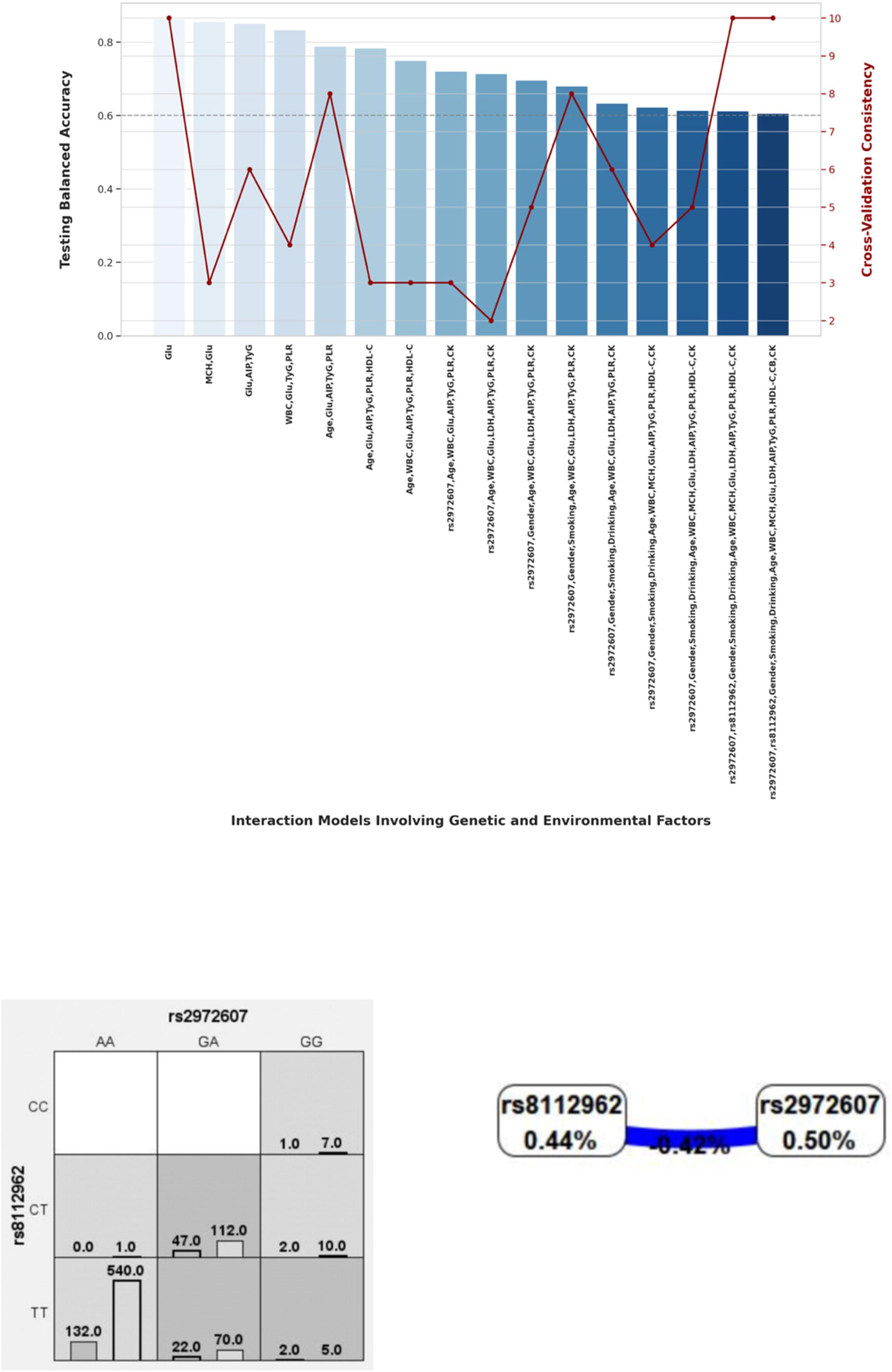
Predictive Performance and Interaction Analysis of MDR Models Involving C5L2 Variants in Comorbid T2DM and CHD. (A) Performance metrics of MDR models incorporating C5L2 variants (rs2972607, rs8112962) and clinical factors (glucose, AIP, TyG, PLR, WBC). Testing balanced accuracy (blue bars) and cross-validation consistency (red line) are shown. All models were statistically significant (P < 0.001); increased model complexity improved training accuracy but reduced testing accuracy, indicating potential overfitting. (B) Genotype combinations of rs8112962 and rs2972607 were analyzed using Multifactor Dimensionality Reduction (MDR). Each cell shows a specific genotype pair, with case (left bar) and control (right bar) counts; shading denotes risk classification (dark grey: high-risk, light grey: low-risk, white: neutral). (C) Interaction dendrogram showing a negative information gain (–0.42%) between rs8112962 and rs2972607, indicating a redundant interaction. Individual SNP contributions are noted (rs8112962: 0.44%, rs2972607: 0.50%).

#### 3.5.2. Gene-Environment Interaction Models

To explore gene-environment interactions in comorbid type 2 diabetes mellitus (T2DM) and coronary heart disease (CHD), MDR analysis was conducted. Figure 7 (Panels A–C) adopts the same color-coding as Figure 6B, classifying genotype–environment combinations into high-risk (dark grey), low-risk (light grey), and neutral (white) groups. These models assess interactions between rs2972607 and rs8112962 with clinical and biochemical markers such as creatine kinase (CK), lactate dehydrogenase (LDH), heart rate, and the triglyceride-glucose (TyG) index. In Figure 7A, rs2972607 interacts with clinical variables, e.g, the GG genotype with elevated CK or TyG is enriched in cases, indicating increased risk. Figure 7B shows similar risk associations for rs8112962. Multivariable models in Figure 7C consistently place rs2972607 in high-risk groupings, reinforcing its role in disease susceptibility. The dendrogram in Figure 7D maps variable interactions, with fasting glucose and TyG forming a strongly redundant cluster (dark blue), suggesting overlapping metabolic information. Inclusion of rs2972607 alters this redundancy, introducing weaker negative or new positive interactions (green/yellow) with WBC, LDH, CK, and HDL-C. In contrast, rs8112962 shows only a weak positive interaction, indicating a subtler effect. Overall, these results underscore the complex gene-environment interplay in cardiometabolic disease, highlighting rs2972607 as a potential mechanistic contributor through both direct and modifying effects on metabolic markers.

**Figure 7.**
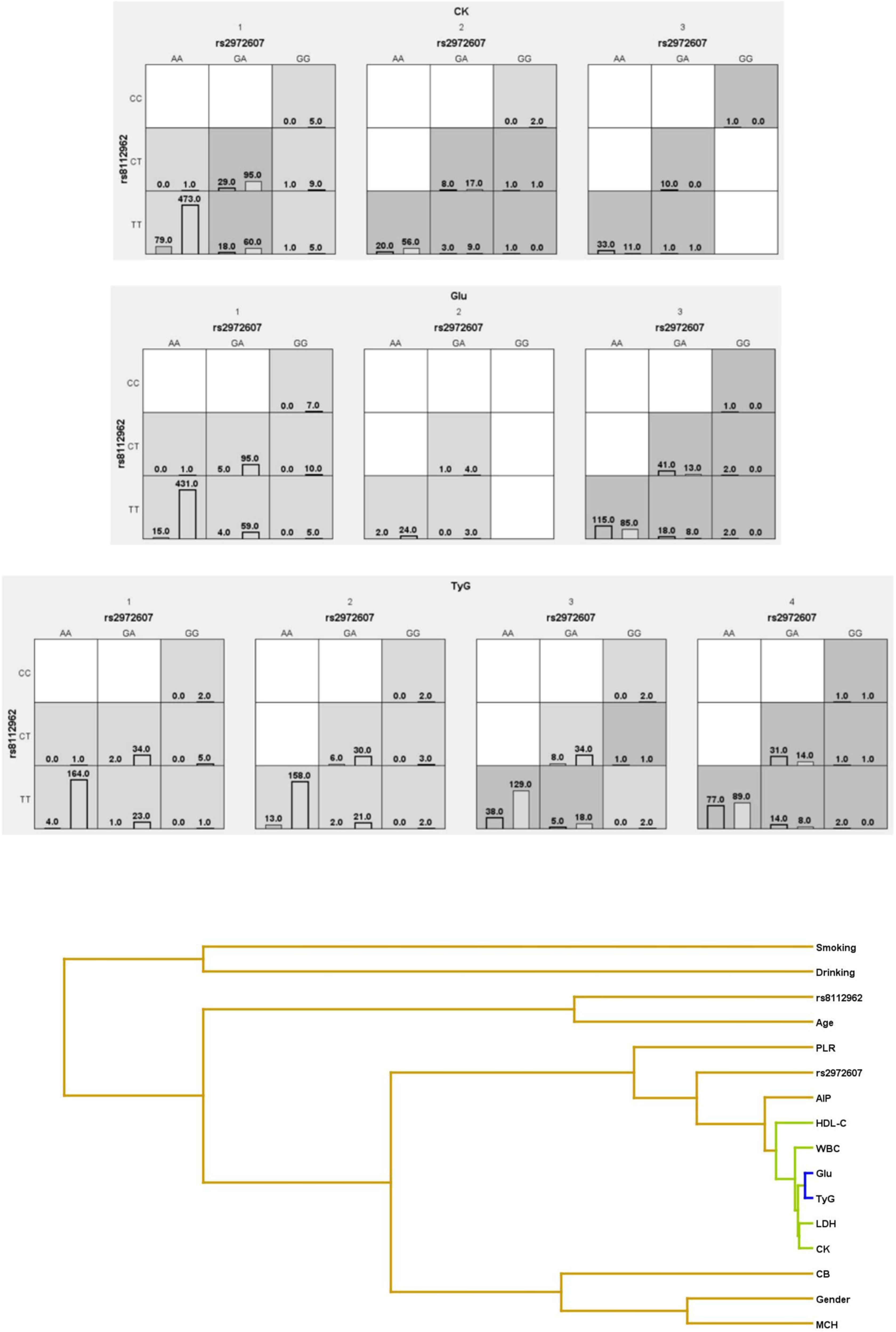
Gene–Environment Interactions Between C5L2 Polymorphisms and Metabolic Biomarkers in Comorbid T2DM and CHD. Multi-dimensional reduction (MDR)-based models reveal gene–environment interactions involving C5L2 polymorphisms (rs2972607, rs8112962) and key clinical biomarkers influencing the risk of comorbid type 2 diabetes mellitus (T2DM) and coronary heart disease (CHD). (A–C) Genotype–biomarker combinations with creatine kinase (CK), fasting glucose (Glu), and triglyceride-glucose (TyG) index are categorized into high-risk (dark grey), low-risk (light grey), and neutral (white) groups. Bars within each cell show the distribution of cases (left) and controls (right). (D) Interaction dendrogram illustrating MDR-based hierarchical relationships; line colors indicate interaction strength: dark blue (strong negative/redundant), green (moderate), yellow (weak positive). (E) Ring network diagram visualizing pairwise interaction effects based on information gain; node size and labels reflect individual main effects, while line thickness and color indicate interaction magnitude and direction (green: positive; blue: negative).

#### 3.5.3. Ring network diagrams of gene-environment interactions

Figure 7E presents the gene–environment interaction network, where each node represents a genetic or environmental factor, labeled with its individual information gain—a measure of its predictive contribution to comorbid T2DM and CHD. Lines between nodes indicate pairwise interactions, with color and thickness denoting their strength and direction. Fasting glucose (Glu) had the highest individual contribution (29.84%), followed by the TyG index (15.81%), CK (7.62%), LDH (7.15%), and WBC (5.78%). Other notable predictors included HDL-C (2.76%), AIP (2.20%), PLR (1.29%), and MCH (0.96%). These clinical markers outperformed demographic and genetic factors. Among genetic variants, C5L2 rs2972607 (0.50%) and rs8112962 (0.44%) showed modest but greater importance than age (0.43%), sex (0.20%), and drinking (0.04%). The strongest negative interaction was between glucose and TyG (–15.81%), likely reflecting metabolic redundancy. The strongest positive interaction was between drinking and smoking (0.18%), highlighting the compounding effect of lifestyle risks. Overall, metabolic factors dominated disease prediction, while genetic variants, though modest individually, may play a more significant role through interactions, supporting integrated gene–environment modeling in understanding complex disease mechanisms.

## 4. Discussion

Coronary heart disease (CHD) and type 2 diabetes mellitus (T2DM) are major global health burdens with overlapping pathophysiological mechanisms. Their frequent co-occurrence heightens morbidity and mortality, yet the genetic and metabolic drivers of this comorbidity remain incompletely understood. This study investigates the role of C5L2 gene polymorphisms (rs2972607 and rs8112962) in T2DM-associated CHD, integrating clinical, inflammatory, and metabolic markers for a holistic risk model. T2DM and CHD share common mechanisms such as insulin resistance, endothelial dysfunction, oxidative stress, and chronic inflammation [6,12,13]. Our analysis identified WBC, LDH, CK, and TyG index as strong predictors of disease risk, emphasizing their role in cardiometabolic pathology. Glucose emerged as the most informative marker, aligning with its central role in diabetic vascular complications. Previous studies also established glucose and TyG index as predictors of CHD severity in T2DM [7,8]; our data extend this by demonstrating interactions with C5L2 variants. C5L2 encodes a G protein-coupled receptor involved in immune and metabolic regulation via ASP and C5a, exerting both pro- and anti-inflammatory effects depending on tissue context [14–22]. While Zheng et al. [11] linked rs2972607 and rs8112962 with CHD in a Chinese Han cohort, we expand on this by characterizing their phenotypic effects in T2DM-CHD comorbidity. We found genotype-specific associations for rs2972607 with LY, PLT, PDW, HDL-C, UCB, AST/ALT ratio, and 5′-NT, and for rs8112962 with monocytes and HDL-C, highlighting their potential roles in lipid metabolism and immune modulation. Our genotype distribution analysis revealed a protective effect of AA (rs2972607) and TT (rs8112962) genotypes, which were underrepresented in cases. This aligns with the concept of genotype-conferred resilience in complex diseases [23,24], and the heterozygote advantage hypothesis [25], with balancing selection possibly maintaining the risk alleles [26]. Multifactor Dimensionality Reduction (MDR) analysis revealed complex gene-gene and gene-environment interactions involving C5L2 SNPs and clinical markers (e.g., WBC, MCH, glucose, LDH, AIP, TyG, PLR, HDL-C, CB, CK). rs2972607 consistently appeared in top MDR models with glucose, WBC, TyG index, and HDL-C, indicating it may influence disease risk via metabolic-inflammatory pathways. The strong negative interaction between glucose and TyG (–15.81%) suggests metabolic redundancy, which rs2972607 modulates in interaction networks, potentially acting as a regulatory hub. In contrast, rs8112962 showed a weaker, more additive effect.

Our integrative strategy is merging genotype–phenotype associations, logistic regression, and high-order interactions, provides a comprehensive framework for dissecting T2DM-CHD comorbidity. This supports the shift away from isolated SNP analysis toward gene–environment and gene–phenotype models. Notably, rs2972607 remained significant after multivariate adjustment, confirming its role as an independent genetic risk factor. These findings align with inflammation-based atherosclerosis models [27–29], positioning C5L2 as a mechanistic link between immune dysregulation and metabolic dysfunction.

However, several limitations warrant caution. The hospital-based, single-center, retrospective design may limit generalizability and introduce bias. The sample size, though adequate for primary outcomes, might lack power for rare variant effects. Future work should employ larger, multi-ethnic, prospective cohorts and functional studies to validate and elucidate the mechanisms of C5L2 polymorphisms in cardiometabolic disease.

In summary, our findings identify C5L2 variants, especially rs2972607, as modulators of T2DM-associated CHD risk through interactions with inflammatory and metabolic pathways. These insights support incorporating genetic screening alongside clinical profiling to enhance risk prediction and guide personalized management of cardiometabolic disease.

## Conclusion

This study advances our understanding of the genetic and metabolic interplay underlying the comorbidity of type 2 diabetes mellitus (T2DM) and coronary heart disease (CHD), highlighting C5L2 gene polymorphisms, particularly rs2972607 as important contributor to disease susceptibility. By integrating clinical biomarkers, logistic regression modeling, and gene–environment interaction analysis, we demonstrate that rs2972607 exerts its influence through complex networks involving metabolic and inflammatory pathways. While rs8112962 showed a more modest role, both variants were found to be associated with specific physiological traits linked to CHD risk in T2DM patients. These findings underscore the importance of evaluating genetic effects in the context of broader biological systems and support the utility of incorporating genetic data into risk prediction frameworks for cardiometabolic diseases.

## Author Contributions

Conceptualization, L.M.; methodology, L.F.; software, D.L.; validation, Y.J.; formal analysis, W.Y.; investigation, Y.M.; resources, J.T.; data curation, L.M.; writing, original draft preparation, L.M. and L.F; writing, review and editing, D.L, Y.J. and W.Y.; visualization, Y.M.; supervision, J.T.; project administration, G.Y.; funding acquisition, G.Y. All authors have read and agreed to the published version of the manuscript.

## Funding

This research was funded by Xinjiang Uygur Autonomous Region Natural Science Foundation, grant number 2023D01D13, and Tianshan Elite Science and Technology Innovation Leading Talents, grant number 2022TSYCLJ0023

## Ethics statement

The Ethical certificate was approved by the ethics committee of The First Affiliated Hospital Medical, Ethics Approval Number: K202309-08, and 240424-14.

## Informed Consent Statement

An Informed Consent for all the subjects was taken according to the ethics documents

## Data Availability Statement

Data are contained within the article. The data presented in this study can be requested from the authors.

## Conflicts of Interest

The authors declare no conflicts of interest.

## Abbreviations

APPT (Activated Partial Thromboplastin Time), PT (Prothrombin Time), BASO (Basophil Count), BAp (Basophil Percentage), CK (Creatine Kinase), CK-MB (Creatine Kinase–MB Isoenzyme), EOS (Eosinophil Count), EOp (Eosinophil Percentage), GGT (Gamma-Glutamyl Transferase), 5′-NT (5′-Nucleotidase), LDH (Lactate Dehydrogenase), MONO (Monocyte Count), MOp (Monocyte Percentage), NEUT (Neutrophil Count), NEp (Neutrophil Percentage), LYp (Lymphocyte Percentage), NLR (Neutrophil-to-Lymphocyte Ratio), PLR (Platelet-to-Lymphocyte Ratio), RBC (Red Blood Cell Count), MCV (Mean Corpuscular Volume), MCH (Mean Corpuscular Hemoglobin), RDW (Red Cell Distribution Width), PDW (Platelet Distribution Width), WBC (White Blood Cell Count), ALT (Alanine Aminotransferase), AST (Aspartate Aminotransferase), A (Albumin), G (Globulin), A/G (Albumin/Globulin Ratio), CB (Conjugated Bilirubin), UCB (Unconjugated Bilirubin), BUN (Blood Urea Nitrogen), UA (Uric Acid), BAR (BUN/Albumin Ratio), Glucose (Blood Glucose), GSP (Glycated Serum Protein), TG (Triglycerides), HDL-C (High-Density Lipoprotein Cholesterol), Apo-A (Apolipoprotein A), LP(a) (Lipoprotein(a)), TyG (Triglyceride-Glucose Index), AIP (Atherogenic Index of Plasma), SII (Systemic Immune-Inflammation Index), SIRI (Systemic Inflammation Response Index), Age (Age in Years), and Heartrate (Heart Rate in Beats per Minute).

## Data Availability

Data are contained within the article

---

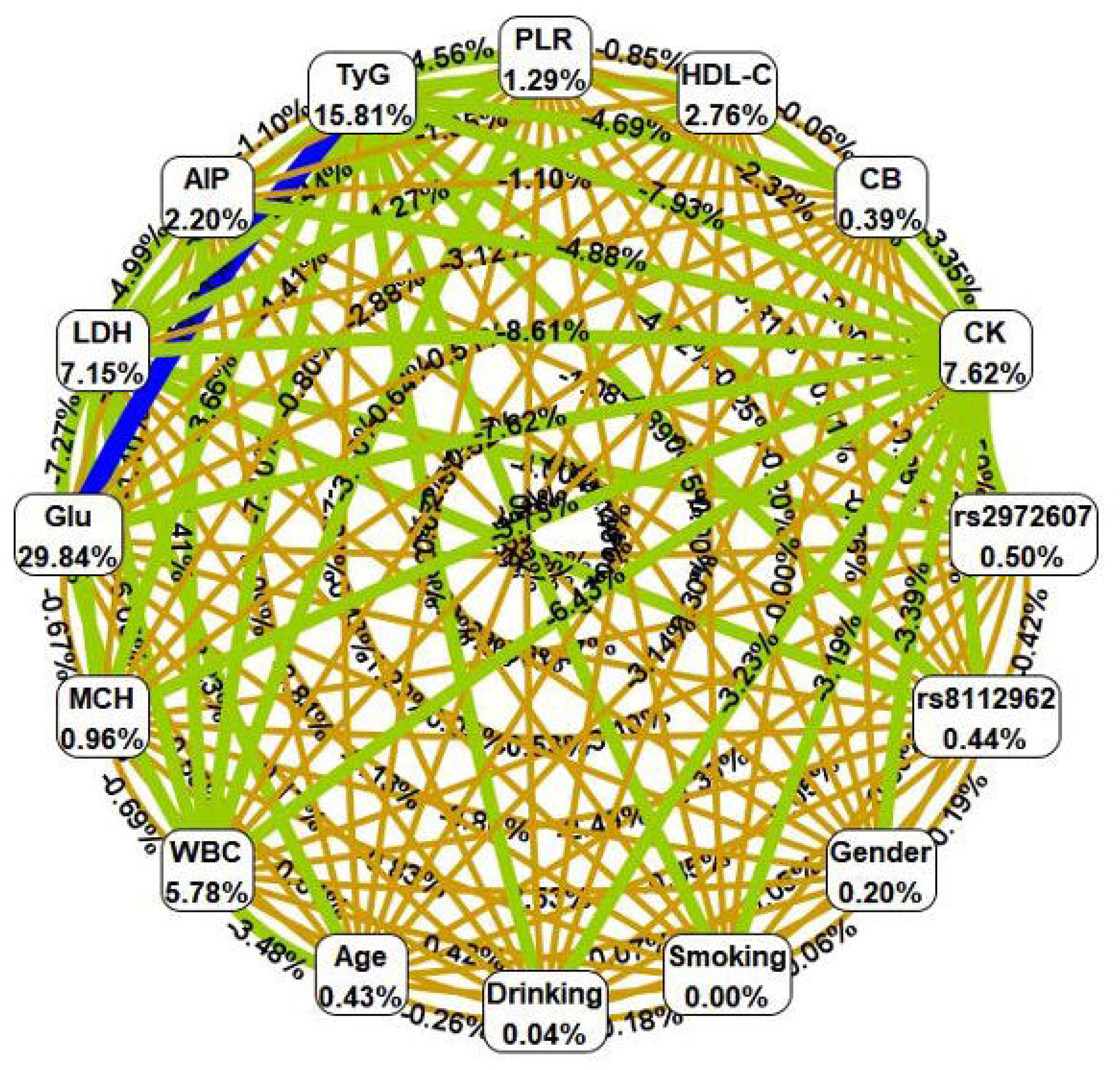

## Notes

### Competing Interest Statement

The authors have declared no competing interest.

### Funding Statement

This study was funded by the Tianshan Elite Science and Technology Innovation Leading Talents grant and the Key Project of Natural Science Foundation of Xinjiang Uygur Autonomous Region grant

### Author Declarations

The study followed the Declaration of Helsinki and was approved by the Ethics Committee of the First Affiliated Hospital of Xinjiang Medical University (Approval No. 240424-14).

