## Supplementary figures for "C5L2 Gene Polymorphisms and Their Interaction with Metabolic and Inflammatory Pathways in T2DM-Associated Coronary Heart Disease: Insights from an Integrative Genetic and Clinical Analysis"

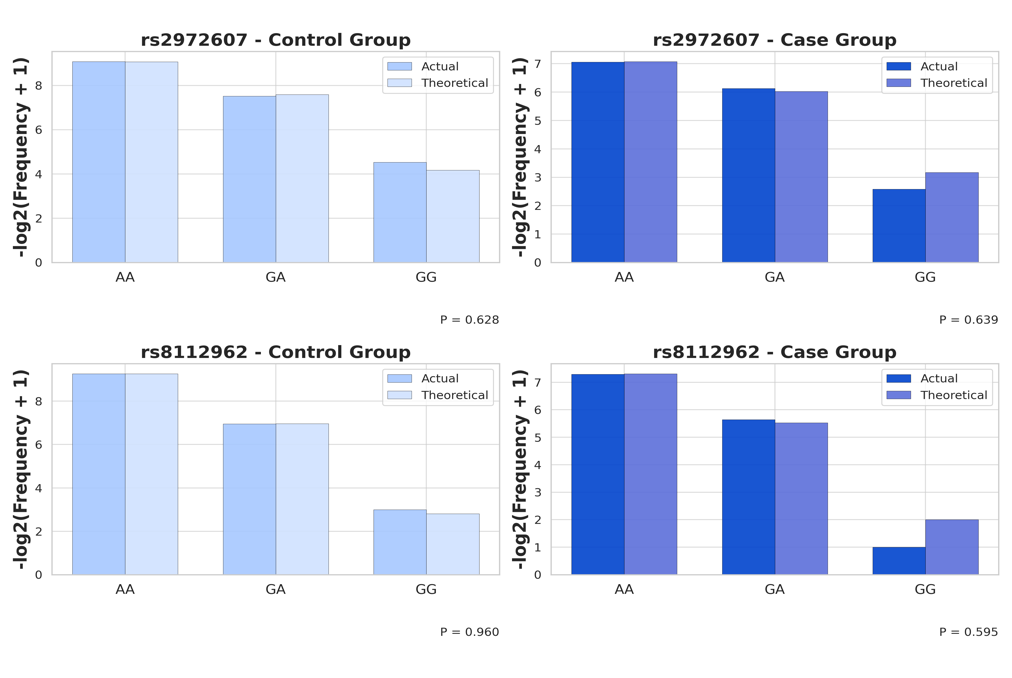

**Supplementary Figure 1. Genotypic Distributions and Hardy-Weinberg Equilibrium Assessment of rs2972607 and rs8112962 in Case and Control Groups.** Bar plots illustrating the actual and theoretical genotype frequencies for the single-nucleotide polymorphisms (SNPs) rs2972607 and rs8112962 in control and case groups. The y-axis represents the log₂-transformed genotype. For each SNP, control groups are depicted in light blue and case groups in dark blue, with actual and Hardy-Weinberg equilibrium (HWE) theoretical values shown side-by-side for each genotype (AA, GA/CT, GG/CC). P-values from the chi-square goodness-of-fit test for HWE are displayed below each subplot’s legend
