## Supplementary Tables for "C5L2 Gene Polymorphisms and Their Interaction with Metabolic and Inflammatory Pathways in T2DM-Associated Coronary Heart Disease: Insights from an Integrative Genetic and Clinical Analysis"

**Supplementary Table 1 Primer list**

| SNP | Sequence（5’-3’） | Product（bp） |
| --- | --- | --- |
| rs2972607 | F: CAGTTGCCACTTAGGAGCATT | 210 |
|  | R: GGGAAGTTATAGCAAAACAGATGGC |  |
| rs8112962 | F: CCTCCTTAAAGCAGGTAGATTGC | 200 |
|  | R: GCAGTCACCAGCCATATGT |  |

Note: F is the forward primer, R is the reverse primer.

**Supplementary Table 2	The baseline characteristics of controls and cases**

| Variables | Control | Case | χ²/Z | P |
| --- | --- | --- | --- | --- |
| n | 745 | 206 |  |  |
| Gender [Male%] | 494(66.3) | 124(60.2) | 2.652 | 0.103 |
| Smoking[n%] | 284(38.1) | 80(38.8) | 0.035 | 0.852 |
| Drinking[n%] | 230(30.9) | 58(28.2) | 0.564 | 0.453 |
| Age [Years (IQR)] | 56(15) | 58(13) | -3.347 | ＜0.001* |
| Heartrate [BPM, M(IQR)] | 76(13) | 80(16) | -4.264 | ＜0.001* |
| WBC [×10^9^/L, M(IQR)] | 6.51(2.11) | 8.69(3.81) | -11.141 | ＜0.001* |
| NEUT [×10^9^/L, M(IQR)] | 3.69(1.69) | 5.375(3.62) | -11.325 | ＜0.001* |
| MONO [×10^9^/L, M(IQR)] | 0.45(0.22) | 0.505(0.3) | -4.419 | ＜0.001* |
| EOS [×10^9^/L, M(IQR)] | 0.13(0.12) | 0.11(0.15) | -3.594 | ＜0.001* |
| BASO [×10^9^/L, M(IQR)] | 0.02(0.03) | 0.04(0.03) | -7.631 | ＜0.001* |
| NEp [%, M(IQR)] | 58(11.31) | 64.78(19.91) | -8.445 | ＜0.001* |
| LYp [%, M(IQR)] | 31.91(10.7) | 25.06(18.34) | -8.484 | ＜0.001* |
| MOp [%, M(IQR)] | 6.81(2.45) | 6.18(3) | -3.562 | ＜0.001* |
| EOp [%, M(IQR)] | 2.13(1.86) | 1.415(1.98) | -6.847 | ＜0.001* |
| BAp [%, M(IQR)] | 0.35(0.3) | 0.41(0.35) | -2.634 | 0.008* |
| LDH [U/L, M(IQR)] | 156.93(39) | 167.325(93.88) | -3.069 | 0.002* |
| CK [U/L, M(IQR)] | 78.2(50.3) | 92.635(183.72) | -3.625 | ＜0.001* |
| CK-MB [U/L, M(IQR)] | 13(6.48) | 18.37(24.26) | -8.604 | ＜0.001* |
| RBC [×10^12^/L, M(IQR)] | 4.66(0.7) | 4.725(0.78) | -2.035 | 0.042* |
| MCV [fl, M(IQR)] | 92.1(5.68) | 90.245(6.13) | -5.149 | ＜0.001* |
| MCH [pg, M(IQR)] | 30.5(2.2) | 30(2.1) | -4.105 | ＜0.001* |
| RDW [CV%, M(IQR)] | 13.1(0.9) | 12.95(0.8) | -3.882 | ＜0.001* |
| PDW [%, M(IQR)] | 16.2(1.72) | 14.34(4.9) | -4.925 | ＜0.001* |
| PT [s, M(IQR)] | 10.8(1.3) | 10.9(1.2) | -2.029 | 0.043* |
| APPT [s, M(IQR)] | 31.1(3.8) | 30.1(4.1) | -3.732 | ＜0.001* |
| BUN [mmol/L, M(IQR)] | 5.3(1.76) | 5.5(2.49) | -2.173 | 0.030* |
| UA [umol/L, M(IQR)] | 304(116.25) | 285.115(108.1) | -3.263 | 0.001* |
| Glucose [mmol/L, M(IQR)] | 4.89(1.02) | 10.72(7.02) | -18.906 | ＜0.001* |
| GSP [mmol/L, M(IQR)] | 2.16(0.4) | 2.5(0.79) | -8.768 | ＜0.001* |
| TG [mmol/L, M(IQR)] | 1.56(1.19) | 1.81(1.36) | -3.68 | ＜0.001 |
| HDL-C [mmol/L, M(IQR)] | 1.06(0.37) | 0.94(0.31) | -6.053 | ＜0.001* |
| Apo-A [g/L, M(IQR)] | 1.19(0.28) | 1.125(0.35) | -2.395 | 0.017* |
| LP(a) [mg/L, M(IQR)] | 147(140.86) | 172.43(210.37) | -2.146 | 0.032* |
| CB [umol/L, M(IQR)] | 3.11(2.14) | 2.225(1.85) | -7.936 | ＜0.001* |
| UCB [umol/L, M(IQR)] | 7.71(5.04) | 8.765(6.59) | -2.754 | 0.006* |
| A [g/L, M(IQR)] | 40.3(4.73) | 39.655(5.27) | -2.507 | 0.012* |
| G [g/L, M(IQR)] | 26(6) | 28.355(6.4) | -5.646 | ＜0.001* |
| A/G[M(IQR)] | 1.55(0.45) | 1.405(0.39) | -6.142 | ＜0.001* |
| AST [U/L, M(IQR)] | 19.5(8.07) | 23.4(27.42) | -5.851 | ＜0.001* |
| ALT [U/L, M(IQR)] | 22(15.61) | 24.17(20.15) | -2.693 | 0.007* |
| GGT [U/L, M(IQR)] | 25.7(21.6) | 29.39(29.9) | -2.158 | 0.031* |
| 5'-NT [U/L, M(IQR)] | 5.3(3.38) | 6.47(5.26) | -5.61 | ＜0.001* |
| AIP [M(IQR)] | 0.16(0.4) | 0.31(0.38) | -5.411 | ＜0.001* |
| SIRI [M(IQR)] | 0.789(0.665) | 1.4845(1.647) | -9.35 | ＜0.001* |
| SII [M(IQR)] | 384.24(246.83) | 606.59(756.06) | -9.089 | ＜0.001* |
| TyG [M(IQR)] | 1.363(0.819) | 2.1815(0.947) | -14.361 | ＜0.001* |
| NLR [M(IQR)] | 1.79(0.98) | 2.62(3.48) | -8.54 | ＜0.001* |
| BAR [M(IQR)] | 0.131(0.049) | 0.141(0.072) | -2.913 | 0.004* |
| PLR [M(IQR)] | 103.6(46.69) | 115.595(68.68) | -2.677 | 0.007* |

Notes：*，statistically significant at P<0.05.

Abbreviations: WBC, white blood cell count; NEUT, neutrophil count; MONO, monocyte count; EOS, eosinophil count; BASO, basophil count; NEp, neutrophil percentage; LYp, lymphocyte percentage; MOp, monocyte percentage; EOp, eosinophil percentage; BAp, basophil percentage; LDH, lactate dehydrogenase; CK, creatine kinase; CK-MB, creatine kinase isoenzyme; RBC, red blood cell count; MCV, mean corpuscular volume of red blood cells; MCH, mean corpuscular hemoglobin volume of red blood cells; RDW, red blood cell distribution width; PDW, platelet distribution width; PT, prothrombin time; APPT, activated partial thromboplastin time; BUN, blood urea nitrogen; UA, uric acid; Glucose, fasting blood glucose; GSP, glycated serum protein; TG, triglyceride; HDL-C, high-density lipoprotein cholesterol; Apo-A, apolipoprotein A; LP(a), lipoprotein (a); CB, bound bilirubin; UCB, unconjugated bilirubin; A, albumin; G, globulin; A/G, albumin/globulin ratio; AST, aspartate aminotransferase; ALT, alanine aminotransferase; GGT, gamma-glutamyl transferase; 5'-NT, 5'-nucleotidase; AIP, plasma atherosclerotic index; SIRI, systemic inflammatory response index; SII, systemic immunoinflammatory index; TyG triglyceride glucose index; NLR, neutrophil/lymphocyte ratio; BAR, basophil/albumin ratio; PLR, platelet/lymphocyte ratio.

**Supplementary Table 3. Hardy Weinberg balance test for two groups of patients**

| **SNP** | **Genotype** | **Control/n (%)** | | | |  | **Case/n (%)** | | | |
| --- | --- | --- | --- | --- | --- | --- | --- | --- | --- | --- |
|  |  | **Actual value** | **Theoretical value** | **χ²** | **P** |  | **Actual value** | **Theoretical value** | **χ²** | **P** |
| rs2972607 | AA | 541(72.62%) | 536(71.95%) | 0.932 | 0.628 |  | 132(64.08%) | 134(65.05%) | 0.895 | 0.639 |
|  | GA | 182(24.43%) | 192(25.77%) |  |  |  | 69(33.50%) | 64(31.07%) |  |  |
|  | GG | 22(2.95%) | 17(2.28%) |  |  |  | 5(2.42%) | 8(3.88%) |  |  |
| rs8112962 | TT | 615(82.55%) | 615(82.55%) | 0.081 | 0.960 |  | 156(75.73%) | 158(76.70%) | 1.122 | 0.595 |
|  | CT | 123(16.51%) | 124(16.64%) |  |  |  | 49(23.79%) | 45(21.84%) |  |  |
|  | CC | 7(0.94%) | 6(0.81%) |  |  |  | 1(0.48%) | 3(1.46%) |  |  |

**Supplementary Table 4. Genotypic distribution of C5L2 polymorphisms rs2972607 and rs8112962 and their statistical comparison between case and healthy controls**

| **SNP** | | **Genotype/Allele** | **Control(*n=*745)** | **Case(*n=*206)** | ***χ*²** | ***P*** |
| --- | --- | --- | --- | --- | --- | --- |
| rs2972607 | Codominant model | AA | 541(72.6%) | 132(64.1%) | 6.854 | 0.033^*^ |
|  |  | GA | 182(24.4%) | 69(33.5%) |  |  |
|  |  | GG | 22(3.0%) | 5(2.4%) |  |  |
|  | Allele | A | 1264(84.8%) | 333(80.8%) | 3.849 | 0.050 |
|  |  | G | 226(15.2%) | 79(19.2%) |  |  |
|  | Dominant model | GG+GA | 204(27.4%) | 74(35.9%) | 5.689 | 0.017^*^ |
|  |  | AA | 541(72.6%) | 132(64.1%) |  |  |
|  | Recessive model | GG | 22(3.0%) | 5(2.4%) | 0.162 | 0.688 |
|  |  | GA+AA | 723(97.0%) | 201(97.6%) |  |  |
|  | Over dominant model | AA+GG | 563(75.6%) | 137(66.5%) | 6.827 | 0.009^*^ |
|  |  | GA | 182(24.4%) | 69(33.5%) |  |  |
|  | Additive model | AA | 541(96.1%) | 132(96.4%) | 0.020 | 0.888 |
|  |  | GG | 22(3.9%) | 5(3.6%) |  |  |
| rs8112962 | Codominant model | TT | 615(82.6%) | 156(75.7%) | 6.046 | 0.049^*^ |
|  |  | CT | 123(16.5%) | 49(23.8%) |  |  |
|  |  | CC | 7(0.9%) | 1(0.5%) |  |  |
|  | Allele | T | 1353(90.8%) | 361(87.6%) | 3.673 | 0.055 |
|  |  | C | 137(9.2%) | 51(12.4%) |  |  |
|  | Dominant model | CT+CC | 130(17.4%) | 50(24.3%) | 4.895 | 0.027^*^ |
|  |  | TT | 615(82.6%) | 156(75.7%) |  |  |
|  | Recessive model | CC | 7(0.9%) | 1(0.5%) | 0.040 | 0.841 |
|  |  | CT+TT | 738(99.1%) | 205(99.5%) |  |  |
|  | Over dominant model | TT+CC | 622(83.5%) | 157(76.2%) | 5.767 | 0.016^*^ |
|  |  | CT | 123(16.5%) | 49(23.8%) |  |  |
|  | Additive model | CC | 7(1.1%) | 1(0.6%) | 0.010 | 0.921 |
|  |  | TT | 615(98.9%) | 156(99.4%) |  |  |

Notes:*, statistically significant at *P*＜0.05.

**Supplementary Table 5 Correlation of C5L2 gene rs2972607 genotypes with clinical** **indicator**

| **Variables** | **rs2972607 Genotype** | | | ***H/χ*²** | ***P*** |
| --- | --- | --- | --- | --- | --- |
|  | AA | GA | GG |  |  |
| *n* | 673 | 251 | 27 |  |  |
| Gender [Male%] | 63.20% | 68.50% | 77.80% | 4.321 | 0.115 |
| Smoking[n%] | 37.6% | 40.2% | 37.00% | 0.560 | 0.756 |
| Drinking[n%] | 30.60% | 29.90% | 25.90% | 0.296 | 0.862 |
| Age [Years, M(IQR)] | 57(15) | 55(14) | 51(16) | 4.132 | 0.127 |
| Weight [Kg, M(IQR)] | 74(16) | 77(16) | 73(20) | 10.623 | 0.005 |
| Breathing [RR, M(IQR)] | 19(2) | 19(2) | 19(2) | 14.266 | ＜0.001* |
| LY [×10^9^/L, M(IQR)] | 2.01(0.8) | 2.09(0.92) | 1.82(0.47) | 7.490 | 0.024* |
| MONO [×10^9^/L, M(IQR)] | 0.46(0.23) | 0.45(0.23) | 0.5(0.28) | 0.349 | 0.84 |
| MOp [%, M(IQR)] | 6.77(2.51) | 6.49(2.93) | 6.86(2.86) | 3.691 | 0.158 |
| PLT [×10^9^/L, M(IQR)] | 215(70) | 219(72) | 188(59) | 8.469 | 0.014* |
| PDW [%, M(IQR)] | 16.02(3.97) | 16.2(2.01) | 16.23(1.56) | 6.146 | 0.046* |
| HDL-C [mmol/L, M(IQR)] | 1.05(0.37) | 0.98(0.36) | 0.95(0.47) | 11.240 | 0.004* |
| UCB [umol/L, M(IQR)] | 8.2(4.98) | 7.38(6.26) | 5.8(4.4) | 8.069 | 0.018* |
| DeRits [M(IQR)] | 0.94(0.52) | 0.89(0.45) | 0.82(0.41) | 8.379 | 0.015* |
| 5'-NT [U/L, M(IQR)] | 5.3(3.59) | 6.1(3.6) | 6.06(5.9) | 8.020 | 0.018* |

Notes:*,statistically significant at P＜0.05.

Abbreviations: LY (lymphocyte count), MONO (monocyte count), MOp (monocyte percentage), PLT (platelet count), PDW (platelet distribution width), HDL-C (high-density lipoprotein cholesterol), UCB (unconjugated bilirubin), DeRits (aspartate aminotransferase/alanine aminotransferase ratio), and 5'-NT (5'-nucleotidase).

**Supplementary Table 6 Correlation of C5L2 gene rs8112962 genotypes with clinical indicators**

| **Variables** | **rs8112962 Genotype** | | | ***H/χ*²** | ***P*** |
| --- | --- | --- | --- | --- | --- |
|  | TT | CT | CC |  |  |
| *n* | 771 | 172 | 8 |  |  |
| Gender [Male%] | 64.50% | 66.30% | 87.50% | 2.002 | 0.368 |
| Smoking[n%] | 38 .30% | 37.20% | 62.50% | 2.049 | 0.397 |
| Drinking[n%] | 30.40% | 29.70% | 37.50% | 0.232 | 0.891 |
| Age [Years, M(IQR)] | 56(14) | 57(16) | 48(12) | 4.783 | 0.092 |
| Weight [Kg, M(IQR)] | 74(17) | 76(17) | 77(15) | 3.507 | 0.173 |
| Breathing [RR, M(IQR)] | 19(2) | 19(2) | 18.5(1) | 6.573 | 0.037* |
| LY [×10^9^/L, M(IQR)] | 2.05(0.79) | 2(0,94) | 2.28(1.21) | 0.629 | 0.730 |
| MONO [×10^9^/L, M(IQR)] | 0.47(0.24) | 0.43(0.26) | 0.49(0.15) | 9.353 | 0.009* |
| Mop [%, M(IQR)] | 6.8(2.53) | 6.24(2.8) | 6.71(2.29) | 10.282 | 0.006* |
| PLT [×10^9^/L, M(IQR)] | 215(70) | 212.5(73) | 176(116) | 2.568 | 0.277 |
| PDW [%, M(IQR)] | 16.1(3.3) | 16.12(2.49) | 16.12(1.9) | 0.603 | 0.740 |
| HDL-C [mmol/L, M(IQR)] | 1.04(0.37) | 0.98(0.38) | 1.14(0.66) | 6.434 | 0.04* |
| UCB [umol/L, M(IQR)] | 8.03(5.01) | 7.44(7.41) | 7.78(3.76) | 1.012 | 0.603 |
| DeRits [M(IQR)] | 0.92(0.52) | 0.9(0.46) | 0.7(0.45) | 2.311 | 0.315 |
| 5^'^-NT [U/L, M(IQR)] | 5.4(3.7) | 6.1(3.68) | 5.78(4.45) | 5.158 | 0.076 |

Notes:*,statistically significant at P＜0.05.

Abbreviations: LY (lymphocyte count), MONO (monocyte count), MOp (monocyte percentage), PLT (platelet count), PDW (platelet distribution width), HDL-C (high-density lipoprotein cholesterol), UCB (unconjugated bilirubin), DeRits (aspartate aminotransferase/alanine aminotransferase ratio), and 5'-NT (5'-nucleotidase).

**Supplementary Table 7 Variable Classification and Coding Scheme for Logistic Regression Analysis**

|  | **Variables** | **Assignment** | |  | **Variables** | **Assignment** | |
| --- | --- | --- | --- | --- | --- | --- | --- |
|  |  | **Man** | **Woman** |  |  | **Man** | **Woman** |
| Y | T2DM+CHD | 0=not | | X26 | LP(a)(mg/L) | 0="＜300.00" | |
|  |  | 1=yes | |  |  | 1="≥300.00" | |
| XI | Gender | 1=Man | | X27 | CB (umol/L) | 0="＜6.80" | |
|  |  | 2=Woman | |  |  | 1="≥6.80" | |
| X2 | Smoking | 0=not | | X28 | UCB (umol/L) | 1="≥1.70&＜10.20" | |
|  |  | 1=yes | |  |  | 2="＜1.70" | |
| X3 | Drinking | 0=not | |  |  | 3="≥10.20" | |
|  |  | 1=yes | | X29 | A(g/L) | 1="≥40.00&＜55.00" | |
| X4 | Age(years) | 1="＜45" | |  |  | 2="＜40.00" | |
|  |  | 2="≥45&＜60" | |  |  | 3="≥55.00" | |
|  |  | 3="≥60" | | X30 | G(g/L) | 1="≥20.00&＜30.00" | |
| X5 | Breathing (RR) | 1="≥12&＜20" | |  |  | 2="＜20.00" | |
|  |  | 2="＜12" | |  |  | 3="≥30.00" | |
|  |  | 3="≥20" | | X31 | A/G | 1="≥1.50&＜2.50" | |
| X6 | Heartrate (BPM) | 1="≥60&＜100" | |  |  | 2="＜1.50" | |
|  |  | 2="＜60" | |  |  | 3="≥2.50" | |
|  |  | 3="≥100" | | X32 | AST(U/L) | 1="≥5.00&＜40.00" | |
| X7 | WBC(×109/L) | 1="≥4.00&＜10.00" | |  |  | 2="＜5.00" | |
|  |  | 2="＜4.00" | |  |  | 3="≥40.00" | |
|  |  | 3="≥10.00" | | X33 | ALT(U/L) | 1="≥8.00&＜40.00" | |
| X8 | NEUT(×109/L) | 1="≥2.00&＜7.00" | |  |  | 2="＜8.00" | |
|  |  | 2="＜2.00" | |  |  | 3="≥40.00" | |
|  |  | 3="≥7.00" | | X34 | GGT(U/L) | 1="≥11.00&＜50.00" | 1="≥7.00&＜32.00" |
| X9 | MONO(×109/L) | 1="≥0.12&＜0.80" | |  |  | 2="＜11.00" | 2="＜7.00" |
|  |  | 2="＜0.12" | |  |  | 3="≥50.00" | 3="≥32.00" |
|  |  | 3="≥0.80" | | X35 | 5'-NT(U/L) | 1="≥1.00&＜11.00" | |
| X10 | LY(×109/L) | 1="≥0.80&＜4.00" | |  |  | 2="＜1.00" | |
|  |  | 2="＜0.80" | |  |  | 3="≥11.00" | |
|  |  | 3="≥4.00" | | X36 | LDH(U/L) | 1="≥120.00&＜250.00" | |
| X11 | EOS(×109/L) | 1="≥0.05&＜0.50" | |  |  | 2="＜120.00" | |
|  |  | 2="＜0.05" | |  |  | 3="≥250.00" | |
|  |  | 3="≥0.50" | | X37 | CK(U/L) | 1="≥50.00&＜310.00" | 1="≥40.00&＜200.00" |
| X12 | BASO(×109/L) | 0="＜0.10" | |  |  | 2="＜50.00" | 2="＜40.00" |
|  |  | 1="≥0.10" | |  |  | 3="≥310.00" | 3="≥200.00" |
| X13 | RBC(×1012/L) | 1="≥4.00&＜5.50" | 1="≥3.50&＜5.00" | X38 | AIP | 1="＜-0.01" | |
|  |  | 2="＜4.00" | 2="＜3.50" |  |  | 2="≥-0.01&＜0.19" | |
|  |  | 3="≥5.50" | 3="≥5.00" |  |  | 3="≥0.19&＜0.40" | |
| X14 | MCV (fl) | 1="≥80.00&＜100.00" | |  |  | 4="≥0.40" | |
|  |  | 2="＜80.00" | | X39 | SIRI | 1="＜-0.561" | |
|  |  | 3="≥100.00" | |  |  | 2="≥-0.561&＜0.877" | |
| X15 | MCH (pg) | 1="≥27.00&＜34.00" | |  |  | 3="≥0.877&＜1.410" | |
|  |  | 2="＜27.00" | |  |  | 4="≥1.410" | |
|  |  | 3="≥34.00" | | X40 | SII | 1="＜288.64" | |
| X16 | RDW (%) | 1="≥11.5&＜14.5" | |  |  | 2="≥288.64&＜407.24" | |
|  |  | 2="＜11.5" | |  |  | 3="≥407.24&＜608.33" | |
|  |  | 3="≥14.5" | |  |  | 4="≥608.33" | |
| X17 | PLT(×109/L) | 1="≥100&＜300" | | X41 | TyG | 1="＜1.061" | |
|  |  | 2="＜100" | |  |  | 2="≥1.061&＜1.523" | |
|  |  | 3="≥300" | |  |  | 3="≥1.523&＜2.027" | |
| X18 | PT(s) | 1="≥11.0&＜14.0" | |  |  | 4="≥2.027" | |
|  |  | 2="＜11.0" | | X42 | NLR | 1="＜1.41" | |
|  |  | 3="≥14.0" | |  |  | 2="≥1.41&＜1.92" | |
| X19 | APTT(s) | 1="≥30.0&＜42.0" | |  |  | 3="≥1.92&＜2.66" | |
|  |  | 2="＜30.0" | |  |  | 4="≥2.66" | |
|  |  | 3="≥42.0" | | X43 | BAR | 1="＜0.109" | |
| X20 | BUN (mmol/L) | 1="≥3.20&＜7.10" | |  |  | 2="≥0.109&＜0.134" | |
|  |  | 2="＜3.20" | |  |  | 3="≥0.134&＜0.161" | |
|  |  | 3="≥7.10" | |  |  | 4="≥0.161" | |
| X21 | UA (umol/L) | 0="＜420.00" | | X44 | PLR | 1="＜84.79" | |
|  |  | 1="≥420.00" | |  |  | 2="≥84.79&＜106.37" | |
| X22 | Glucose(mmol/L) | 1="≥3.90&＜6.10" | |  |  | 3="≥106.37&＜134.09" | |
|  |  | 2="＜3.90" | |  |  | 4="≥134.09" | |
|  |  | 3="≥6.10" | | X45 | rs2972607 | 1=AA | |
| X23 | TG (mmol/L) | 0="＜2.30" | |  |  | 2=GA | |
|  |  | 1="≥2.30" | |  |  | 3=GG | |
| X24 | HDL-C(mmol/L) | 0="＞1.00" | | X46 | rs8112962 | 1=TT | |
|  |  | 1="≤1.00" | |  |  | 2=CT | |
| X25 | Apo-A(g/L) | 1="≥1.25&＜1.59" | 1="≥1.31&＜1.59" |  |  | 3=CC | |
|  |  | 2="＜1.25" | 2="＜1.31" |  |  |  | |
|  |  | 3="≥1.59" | 3="≥1.59" |  |  |  | |

Note: This table presents the categorical assignment of clinical, biochemical, and genetic variables used in logistic regression models to evaluate risk factors associated with comorbid type 2 diabetes mellitus (T2DM) and coronary heart disease (CHD). Each variable was stratified into clinically meaningful categories, with corresponding numeric codes applied for regression analysis. Genetic variants rs2972607 and rs8112962 of the **C5L2** gene were coded by genotype. Y denotes the outcome variable indicating T2DM+CHD status (0 = no, 1 = yes). **Abbreviations**: WBC (white blood cell count), NEUT (neutrophil count), MONO (monocyte count), LY (lymphocyte count), EOS (eosinophil count), and BASO (basophil count) refer to key leukocyte subsets. RBC (red blood cell count), MCV (mean corpuscular volume of red blood cells), MCH (mean corpuscular hemoglobin content of red blood cells), and RDW (red blood cell distribution width) are hematological parameters related to erythrocyte morphology and function. PLT (platelet count), PT (prothrombin time), and APTT (activated partial thromboplastin time) are indicators of coagulation. BUN (blood urea nitrogen), UA (uric acid), and Glucose (fasting glucose) are metabolic markers, while TG (triglyceride), HDL-C (high-density lipoprotein cholesterol), Apo-A (apolipoprotein A), and LP(a) [lipoprotein (a)] are lipid-related indicators. CB (bound bilirubin), UCB (unconjugated bilirubin), A (albumin), G (globulin), and A/G (albumin/globulin ratio) reflect liver and protein metabolism. Liver enzymes include AST (aspartate aminotransferase), ALT (alanine aminotransferase), GGT (gamma-glutamyl transferase), and 5'-NT (5'-nucleotidase), while LDH (lactate dehydrogenase) and CK (creatine kinase) are markers of tissue damage and cellular turnover. Composite indices include AIP (atherosclerotic index of plasma), SIRI (systemic inflammatory response index), SII (systemic immune response index), TyG (triglyceride glucose index), NLR (neutrophil/lymphocyte ratio), BAR (basophil/albumin ratio), and PLR (platelet/lymphocyte ratio), which capture systemic inflammation, immune balance, and metabolic status.

**Supplementary Table 8. Univariate Logistic Regression Results for Clinical, Biochemical, and Genetic Predictors of T2DM with CHD**

| **Variables** | **P value** | **OR** | **95% CI for OR** | |  | **Variables** | **P value** | **OR** | **95% CI for OR** | |
| --- | --- | --- | --- | --- | --- | --- | --- | --- | --- | --- |
|  |  |  | **Lower** | **Upper** |  |  |  |  | **Lower** | **Upper** |
| Breathing | 0.015* | 1.485 | 1.081 | 2.041 |  | A/G | ＜0.001* |  |  |  |
| Heartrate | 0.002* |  |  |  |  | A/G (2) | ＜0.001* | 2.325 | 1.688 | 3.202 |
| Heartrate (2) | 0.001* | 0.296 | 0.148 | 0.593 |  | A/G (3) | 0.844 | 0.809 | 0.098 | 6.671 |
| Heartrate (3) | 0.013* | 0.225 | 0.070 | 0.727 |  | AST (2) | ＜0.001* | 6.758 | 4.330 | 10.548 |
| WBC | ＜0.001* |  |  |  |  | ALT | 0.068 |  |  |  |
| WBC (2) | 0.079 | 0.167 | 0.022 | 1.234 |  | ALT (2) | 0.999 | 0.000 | 0.000 | . |
| WBC (3) | ＜0.001* | 5.784 | 3.823 | 8.750 |  | ALT (3) | 0.02* | 1.595 | 1.075 | 2.367 |
| NEUT | ＜0.001* |  |  |  |  | GGT | 0.019* |  |  |  |
| NEUT (2) | 0.064 | 0.152 | 0.021 | 1.117 |  | GGT (2) | 0.868 | 1.100 | 0.359 | 3.365 |
| NEUT (3) | ＜0.001* | 9.459 | 6.097 | m |  | GGT (3) | 0.005* | 1.639 | 1.162 | 2.312 |
| MONO | ＜0.001* |  |  |  |  | 5'-NT | ＜0.001* |  |  |  |
| MONO (2) | 0.022* | 3.625 | 1.202 | 10.927 |  | 5'-NT (2) | 0.999 | 0.000 | 0.000 | . |
| MONO (3) | ＜0.001* | 3.994 | 2.427 | 6.574 |  | 5'-NT (3) | ＜0.001* | 2.979 | 1.916 | 4.631 |
| LY | ＜0.001* |  |  |  |  | LDH | ＜0.001 |  |  |  |
| LY (2) | ＜0.001* | 5.653 | 2.380 | 13.427 |  | LDH (2) | 0.024* | 1.801 | 1.079 | 3.007 |
| LY (3) | 0.039* | 2.609 | 1.051 | 6.475 |  | LDH (3) | ＜0.001* | 16.806 | 8.828 | 31.993 |
| EOS | ＜0.001* |  |  |  |  | CK | ＜0.001* |  |  |  |
| EOS (2) | ＜0.001* | 2.803 | 1.833 | 4.285 |  | CK (2) | 0.003* | 1.965 | 1.260 | 3.065 |
| EOS (3) | 0.252 | 0.493 | 0.147 | 1.652 |  | CK (3) | ＜0.001* | 18.984 | 9.769 | 36.893 |
| BASO | 0.038* | 2.877 | 1.058 | 7.820 |  | AIP | ＜0.001* |  |  |  |
| RBC | ＜0.001* |  |  |  |  | AIP (2) | 0.011* | 1.948 | 1.168 | 3.248 |
| RBC (3) | ＜0.001* | 2.834 | 1.696 | 4.737 |  | AIP (4) | ＜0.001* | 3.497 | 2.156 | 5.671 |
| MCH | 0.006* |  |  |  |  | SIRI | ＜0.001* |  |  |  |
| MCH (3) | 0.066 | 0.152 | 0.020 | 1.132 |  | SIRI (3) | ＜0.001* | 5.638 | 3.892 | 8.168 |
| PLT | 0.014* |  |  |  |  | SII | ＜0.001* |  |  |  |
| PLT (2) | 0.099 | 3.867 | 0.774 | 19.319 |  | SII (2) | 0.689 | 1.118 | 0.648 | 1.930 |
| PLT (3) | 0.014* | 1.933 | 1.142 | 3.274 |  | SII (3) | 0.044* | 1.693 | 1.014 | 2.826 |
| APTT | ＜0.001* |  |  |  |  | SII (4) | ＜0.001* | 5.695 | 3.558 | 9.116 |
| APTT (2) | ＜0.001* | 1.878 | 1.368 | 2.578 |  | TyG | ＜0.001* |  |  |  |
| APTT (3) | 0.048* | 2.589 | 1.007 | 6.654 |  | TyG (2) | 0.009* | 3.194 | 1.331 | 7.666 |
| BUN (2) | 0.965 | 1.018 | 0.460 | 2.253 |  | TyG (4) | ＜0.001* | 36.637 | 16.566 | 81.026 |
| BUN (3) | ＜0.001* | 2.083 | 1.391 | 3.119 |  | NLR | ＜0.001* |  |  |  |
| UA | 0.032* | 0.537 | 0.305 | 0.948 |  | NLR (2) | 0.492 | 0.829 | 0.486 | 1.416 |
| Glucose | ＜0.001* |  |  |  |  | NLR (3) | 0.242 | 1.343 | 0.819 | 2.203 |
| Glucose (2) | 0.161 | 2.452 | 0.700 | 8.585 |  | NLR (4) | ＜0.001* | 4.404 | 2.812 | 6.897 |
| Glucose (3) | ＜0.001* | 42.780 | 26.648 | 68.679 |  | BAR | ＜0.001* |  |  |  |
| TG | 0.002* | 1.702 | 1.218 | 2.378 |  | BAR (2) | 0.265 | 0.770 | 0.487 | 1.219 |
| HDL-C | ＜0.001* | 2.638 | 1.910 | 3.643 |  | BAR (3) | 0.170 | 0.721 | 0.452 | 1.151 |
| LP(a) | 0.002* | 1.812 | 1.250 | 2.627 |  | BAR (4) | 0.009* | 1.736 | 1.149 | 2.624 |
| CB | 0.047* | 0.349 | 0.123 | 0.987 |  | PLR | 0.001* |  |  |  |
| UCB | 0.006* |  |  |  |  | PLR (2) | 0.012* | 0.539 | 0.333 | 0.873 |
| UCB (2) | 0.059 | 3.077 | 0.960 | 9.863 |  | PLR (3) | 0.980 | 0.995 | 0.646 | 1.532 |
| UCB (3) | 0.006* | 1.579 | 1.143 | 2.181 |  | PLR (4) | 0.121 | 1.389 | 0.917 | 2.104 |
| G | ＜0.001* |  |  |  |  | rs2972607 | 0.034* |  |  |  |
| G (2) | 0.029* | 0.315 | 0.112 | 0.888 |  | rs2972607(2) | 0.01* | 1.554 | 1.110 | 2.175 |
| G (3) | 0.001* | 1.785 | 1.265 | 2.517 |  | rs2972607(3) | 0.888 | 0.931 | 0.346 | 2.506 |

Notes:*, statistically significant at P＜0.05. **Abbreviations**: WBC (white blood cell count), NEUT (neutrophil count), MONO (monocyte count), LY (lymphocyte count), EOS (eosinophil count), and BASO (basophil count) refer to key leukocyte subsets. RBC (red blood cell count), MCV (mean corpuscular volume of red blood cells), MCH (mean corpuscular hemoglobin content of red blood cells), and RDW (red blood cell distribution width) are hematological parameters related to erythrocyte morphology and function. PLT (platelet count), PT (prothrombin time), and APTT (activated partial thromboplastin time) are indicators of coagulation. BUN (blood urea nitrogen), UA (uric acid), and Glucose (fasting glucose) are metabolic markers, while TG (triglyceride), HDL-C (high-density lipoprotein cholesterol), Apo-A (apolipoprotein A), and LP(a) [lipoprotein (a)] are lipid-related indicators. CB (bound bilirubin), UCB (unconjugated bilirubin), A (albumin), G (globulin), and A/G (albumin/globulin ratio) reflect liver and protein metabolism. Liver enzymes include AST (aspartate aminotransferase), ALT (alanine aminotransferase), GGT (gamma-glutamyl transferase), and 5'-NT (5'-nucleotidase), while LDH (lactate dehydrogenase) and CK (creatine kinase) are markers of tissue damage and cellular turnover. Composite indices include AIP (atherosclerotic index of plasma), SIRI (systemic inflammatory response index), SII (systemic immune response index), TyG (triglyceride glucose index), NLR (neutrophil/lymphocyte ratio), BAR (basophil/albumin ratio), and PLR (platelet/lymphocyte ratio), which capture systemic inflammation, immune balance, and metabolic status.

**Supplementary Table 9 Multivariate Logistic Regression Analysis of Risk Factors for T2DM with CHD**

| **Variables** | **P value** | **OR** | **95% CI for OR** | |
| --- | --- | --- | --- | --- |
|  |  |  | **Lower** | **Upper** |
| WBC | 0.003* |  |  |  |
| WBC (2) | 0.341 | 0.317 | 0.030 | 3.375 |
| WBC (3) | 0.001* | 3.043 | 1.553 | 5.963 |
| MCH | 0.019* |  |  |  |
| MCH (2) | 0.012* | 4.425 | 1.393 | 14.057 |
| MCH (3) | 0.210 | 0.118 | 0.004 | 3.348 |
| Glucose | ＜0.001* |  |  |  |
| Glucose (2) | 0.027* | 4.935 | 1.194 | 20.394 |
| Glucose (3) | ＜0.001* | 14.082 | 7.666 | 25.870 |
| HDL-C | ＜0.001* | 3.765 | 2.002 | 7.079 |
| CB | 0.016* | 0.114 | 0.020 | 0.662 |
| LDH | 0.001* |  |  |  |
| LDH (2) | 0.256 | 0.656 | 0.317 | 1.357 |
| LDH (3) | 0.001* | 8.091 | 2.434 | 26.893 |
| CK | 0.042* |  |  |  |
| CK (2) | 0.024* | 2.127 | 1.107 | 4.088 |
| CK (3) | 0.202* | 2.012 | 0.687 | 5.895 |
| AIP | 0.006* |  |  |  |
| AIP (2) | 0.012* | 0.278 | 0.103 | 0.754 |
| AIP (3) | 0.001* | 0.165 | 0.055 | 0.494 |
| AIP (4) | 0.001* | 0.097 | 0.026 | 0.365 |
| TyG | ＜0.001* |  |  |  |
| TyG (2) | 0.053 | 3.193 | 0.984 | 10.358 |
| TyG (3) | ＜0.001* | 12.003 | 3.215 | 44.805 |
| TyG (4) | ＜0.001* | 32.178 | 7.265 | 142.527 |
| PLR | 0.041* |  |  |  |
| PLR (2) | 0.005* | 0.368 | 0.183 | 0.737 |
| PLR (3) | 0.301 | 0.702 | 0.360 | 1.371 |
| PLR (4) | 0.424 | 0.764 | 0.396 | 1.476 |
| rs2972607 | 0.021* |  |  |  |
| rs2972607(2) | 0.007* | 2.066 | 1.219 | 3.503 |
| rs2972607(3) | 0.289 | 2.352 | 0.484 | 11.438 |

Notes:*,statistically significant at P＜0.05. **Abbreviations**: WBC (white blood cell count), MCH (mean corpuscular hemoglobin content of red blood cells), Glucose (fasting glucose), HDL-C (high-density lipoprotein cholesterol), CB (bound bilirubin), LDH (lactate dehydrogenase), CK (creatine kinase), AIP (atherogenic index of plasma), TyG (triglyceride-glucose index), and PLR (platelet-lymphocyte ratio).

**Supplementary Table 10. Multifactor Dimensionality Reduction (MDR) Models Assessing Gene–Environment Interactions Involving *C5L2* Polymorphisms in T2DM with CHD**

| **Model** | **Training Bal.Acc.CV** | **Testing Bal.Acc.CV** | **CV Consistency** | **P value** |
| --- | --- | --- | --- | --- |
| Glu | 0.8633 | 0.8633 | 10/10 | ＜0.001* |
| MCH, Glu | 0.8663 | 0.8562 | 3/10 | ＜0.001* |
| Glu, AIP, TyG | 0.8744 | 0.8507 | 6/10 | ＜0.001* |
| WBC, Glu, TyG, PLR | 0.8873 | 0.8335 | 4/10 | ＜0.001* |
| Age, Glu, AIP, TyG, PLR | 0.9081 | 0.7889 | 8/10 | ＜0.001* |
| Age, Glu, AIP, TyG, PLR, HDL-C | 0.9278 | 0.7842 | 3/10 | ＜0.001* |
| Age, WBC, Glu, AIP, TyG, PLR, HDL-C | 0.9466 | 0.7496 | 3/10 | ＜0.001* |
| rs2972607, Age, WBC, Glu, AIP, TyG, PLR, CK | 0.9628 | 0.7209 | 3/10 | ＜0.001* |
| rs2972607, Age, WBC, Glu, LDH, AIP, TyG, PLR, CK | 0.9753 | 0.7145 | 2/10 | ＜0.001* |
| rs2972607, Gender, Age, WBC, Glu, LDH, AIP, TyG, PLR, CK | 0.9841 | 0.6966 | 5/10 | ＜0.001* |
| rs2972607, Gender, Smoking, Age, WBC, Glu, LDH, AIP, TyG, PLR, CK | 0.9893 | 0.68 | 8/10 | ＜0.001* |
| rs2972607, Gender, Smoking, Drinking, Age, WBC, Glu, LDH, AIP, TyG, PLR, CK | 0.9924 | 0.6334 | 6/10 | ＜0.001* |
| rs2972607, Gender, Smoking, Drinking, Age, WBC, MCH, Glu, AIP, TyG, PLR, HDL-C, CK | 0.994 | 0.6231 | 4/10 | ＜0.001* |
| rs2972607, Gender, Smoking, Drinking, Age, WBC, MCH, Glu, LDH, AIP, TyG, PLR, HDL-C, CK | 0.9952 | 0.6139 | 5/10 | ＜0.001* |
| rs2972607, rs8112962, Gender, Smoking, Drinking, Age, WBC, MCH, Glu, LDH, AIP, TyG, PLR, HDL-C, CK | 0.9957 | 0.6124 | 10/10 | ＜0.001* |
| rs2972607, rs8112962, Gender, Smoking, Drinking, Age, WBC, MCH, Glu, LDH, AIP, TyG, PLR, HDL-C, CB, CK | 0.9957 | 0.6057 | 10/10 | ＜0.001* |

Notes:*,statistically significant at P＜0.05.
